# Genome-wide analyses identify 147 loci associated with obstructive sleep apnea

**DOI:** 10.1101/2025.11.08.25339824

**Authors:** Luis M. García-Marín, Zuriel Ceja, Santiago Díaz-Torres, Abishna Parasuraman, Jia Wen Xu, Victor Flores-Ocampo, Asma M. Aman, Bade Uckac, Mateo Maya-Martínez, Xueyan Huang, Camilla Pasquali, Aura Aguilar-Roldán, Fangyuan Cao, Natalia S. Ogonowski, Dan J. Siskind, Penelope A. Lind, Ian B. Hickie, Enda M. Byrne, Sarah E. Medland, Nicholas G. Martin, Stuart MacGregor, Xianjun Dong, Sarah J. Lewis, Mathias Seviiri, Jiao Wang, Miguel E. Rentería

**Author notes:** Correspondence: Miguel E. Rentería.

## Abstract

The genetic components underlying obstructive sleep apnoea (OSA) remain poorly understood. Here, we conducted the largest GWAS meta-analysis (OSA; N_cases_= 232,292; N_controls_= 1,259,390) using European ancestry genetic data from five countries. We identified 147 independent loci and estimated SNP-based heritability at 16%. We report six independent loci in a separate African population meta-analysis (N_cases_= 46,834; N_controls_= 149,192). Spatially resolved gene enrichment in neural circuits involved synaptic transmission, including GABAergic and glutamatergic signalling, and cytoskeletal remodelling. OSA-derived polygenic risk scores predicted clinician-ascertained OSA status, Fitbit-derived sleep features, and symptom-based OSA risk and snoring in participants of diverse ancestral backgrounds. We identified putative causal relationships with ADHD, depression, multisite chronic pain, body mass index, and schizophrenia, among others. Our findings demonstrate a substantial genetic component underlying OSA risk beyond shared genetic effects with body mass index, implicating neurobiological pathways related to synaptic function and corticothalamic feedback loops.

## INTRODUCTION

Obstructive sleep apnoea (OSA) is characterised by pharyngeal narrowing and upper airway collapse during sleep, resulting in halted breathing and fragmented non-restorative sleep^1^. Its most prominent symptoms include loud snoring, and gasping for air^1^. Although frequently underdiagnosed, OSA is a common sleep-related disturbance affecting an estimated 1 billion people worldwide^2^. Its prevalence is notably higher in males (25-30%) than in females (9-17%)^2–4^.

The aetiology of OSA is multifactorial, with neuromuscular and lifestyle factors, reduced ventilatory drive during sleep, and individual anatomy (e.g., tongue size and position) all considered important contributors^1^. Among these, obesity, typically measured by the body mass index (BMI), is a well-established major risk factor^5^, with genetic studies suggesting the genetic risk for OSA is partly mediated by obesity^6–8^. While twin and family studies indicate that OSA is heritable (25-87%)^6,7,9^, and genome-wide association studies (GWAS) have identified up to 49 risk loci^6,7^, a comprehensive understanding of its genetic architecture independent of obesity remains incomplete.

In the present study, we analysed individual- and summary-level genetic data for OSA from 1.4 million participants across eight international cohorts. We conducted a large-scale GWAS meta-analysis in individuals of European ancestry, followed by comprehensive post-GWAS analyses. These included fine-mapping, gene-based tests, expression quantitative trait loci (eQTL) mapping, and the integration of spatial transcriptomics data to investigate the putative role of specific brain structures in OSA^10,11^. We also evaluated the predictive utility of OSA-derived polygenic risk scores (PRS) in individuals from diverse ancestral backgrounds. Finally, we explored the genetic overlap and investigated putative causal relationships between OSA and a range of physical and neuropsychiatric phenotypes, sleep disturbances, lifestyle factors, and structural brain neuroimaging measures. This work advances our understanding of the molecular underpinnings of OSA, elucidating its relationships with physical and mental health while providing evidence for its genetic components beyond the effects of BMI.

## RESULTS

### Discovery Genome-Wide Association Analyses in Europeans

Our meta-analysis of European-ancestry GWAS summary data for OSA (N_cases_= 232,292; N_controls_= 1,259,390; **Table 1**) identified 147 independent genome-wide significant loci (*P* < 5 × 10^−8^) (**Figure 1** and **Supplementary Tables 1 and 2**), with the strongest association observed for SNP rs1421085 in chromosome 16 within the *FTO* gene. The single-nucleotide polymorphism (SNP)-based heritability on the liability scale was estimated at 16.26% (SE = 0.006). The genomic inflation factor for the meta-analysis was 1.51. The linkage disequilibrium (LD) score regression intercept was 1.005 (SE = 0.01), with an attenuation ratio of 0.009 (SE = 0.01), suggesting that confounding from population stratification was negligible.

**Figure 1.**
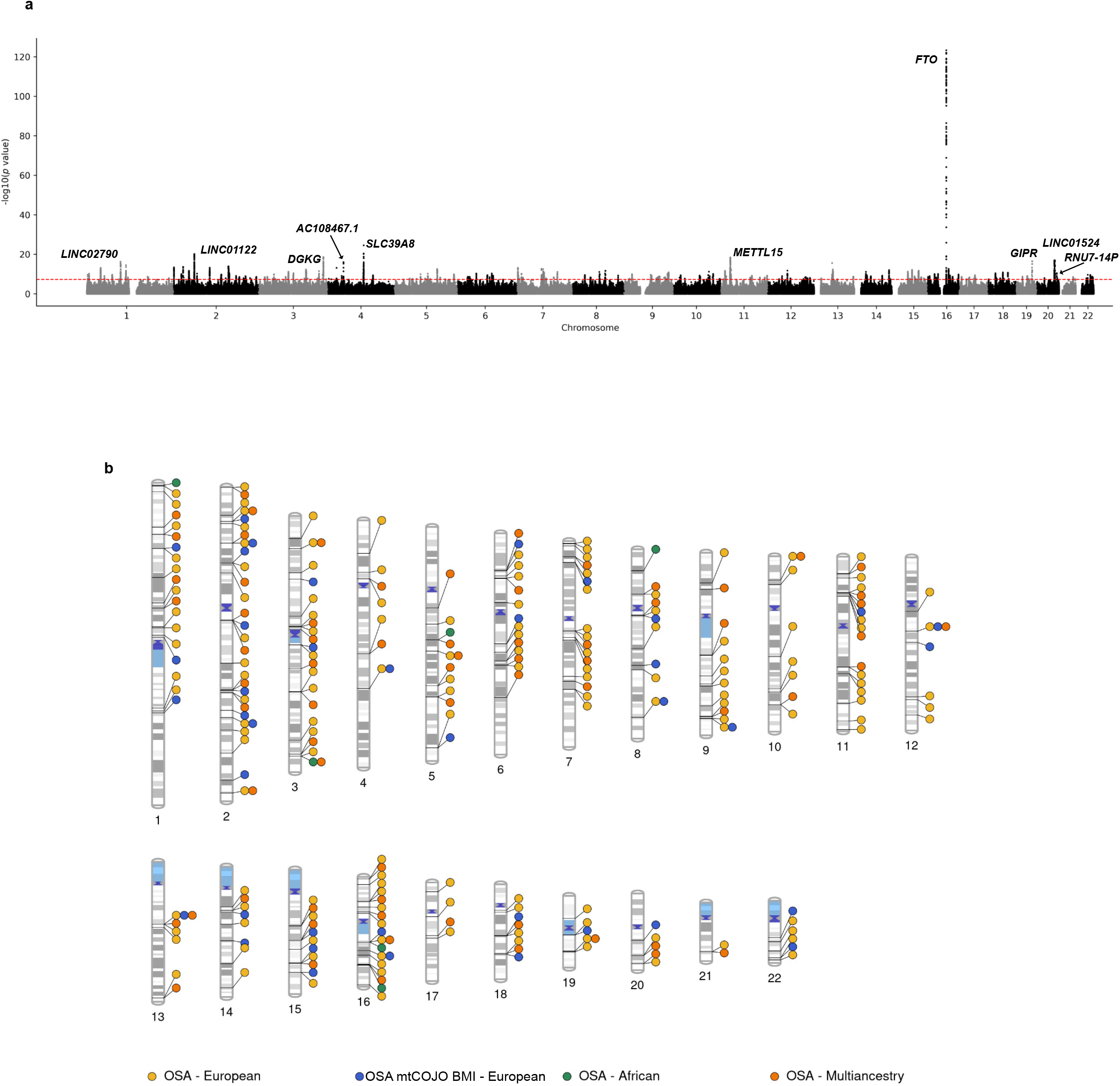
Genome-wide association results for obstructive sleep apnoea (OSA). a) Manhattan plot of the European-ancestry OSA meta-analysis. Each point represents a single nucleotide polymorphism (SNP), plotted by chromosomal position (x-axis) and −log_10_(P) association p-value (y-axis). The red dashed line indicates the genome-wide significance threshold (P < 5 × 10^−08^). Gene names are shown for the top 10 genome-wide significant loci ranked by association strength based on their p-value. Association p-values correspond to two-sided tests from the meta-analysis implemented in METAL (b) Phenogram showing independent genome-wide significant loci across meta-analyses. Lead variants are displayed at their chromosomal positions for the European OSA meta-analysis (yellow), European OSA_mtCOJO_BMI_ (blue), African-ancestry meta-analysis (green), and multi-ancestry meta-analysis (orange). Loci are defined based on LD-based clumping as described in Methods.

**Table 1.** Cohort overview table for European ancestry meta-analysis.

| Cohort | GWAS Conducted or Downloaded | OSA Total Sample Size | OSA Cases | OSA Controls | Phenotype Ascertainment | Phenotype Definition |
|---|---|---|---|---|---|---|
| UK Biobank | Conducted | 270,279 | 5,959 | 264,320 | Clinically ascertained + self-reported | ICD-10 sleep apnoea diagnosis code (code G47.3 in the 41270 UKB data field), and self-reported sleep apnoea (code 1123 in the 20002 UKB data field). Participants with only ‘Yes’ or ‘No’ responses were included, leaving out those who answered 'I don’t know' or 'Prefer not to answer'. Participants who self-reported snoring (defined using data field 1210) were removed from the controls in the OSA GWAS due to the possibility of representing undiagnosed OSA. |
| Million Veteran Program (MVP) | Downloaded | 430,325 | 126,695 | 303,630 | Clinically ascertained | Electronic health records using a multimodal automated phenotyping procedure, which analyses ICD codes and natural language processing of clinical notes. |
| FinnGen | Downloaded | 520,210 | 58,520 | 461,690 | Clinically ascertained | ICD-10 diagnosis code G47.3 (sleep apnoea), corresponding to the FinnGen endpoint G6_SLEEPAPNO. |
| All of Us | Conducted | 225,598 | 32,618 | 192,980 | Clinically ascertained | SNOMED vocabulary concept ID 78275009. Participants who self-reported snoring were removed from the controls in the OSA GWAS due to the possibility of representing undiagnosed OSA. |
| Canadian Longitudinal Study on Aging (CLSA) | Conducted | 15,587 | 4,151 | 11,436 | Self-reported | Via the categories “SNO_STOPBREATH_MCQ” and “SNO_STOPBREATH_COF1” from baseline and follow-up surveys. These categories include the item 'stop breathing in sleep', with ‘Yes’, ‘No’, and 'Don’t know' as possible responses. Participants who responded ‘Don’t know’ were excluded. If a participant reported 'Yes' at baseline but 'No' in the follow-up survey or vice versa, they were deemed as cases. Participants who self-reported snoring were removed from the controls in the OSA GWAS. |
| Australian Cohorts (AGDS & GBP) | Conducted | 9,636 | 1,247 | 8,389 | Self-reported | Using two items: 'During the last month, on how many nights or days per week have you had or been told you had snorting or gasping?’ and ‘During the last month, on how many nights or days per week have you had or been told you had your breathing stop or you choked or struggled for breath?’ Participants were classified as OSA cases if they reported a frequency anywhere between 1 to 7 times per week for either item and were deemed snorers. Controls included individuals who did not snore or have OSA. |
| Mass General Brigham Biobank | Downloaded | 20,047 | 3,102 | 16,945 | Clinically ascertained | Electronic health records data and the ICD-10 sleep apnoea diagnosis code (G47.3). |
| Meta-analysis total | Conducted | 1,491,682 | 232,292 | 1,259,390 | — | — |

All cohorts and sample sizes shown correspond to participants of European genetic ancestry included in the primary OSA meta-analysis. “GWAS Conducted or Downloaded” indicates whether the genome-wide association analysis (GWAS) was newly performed by the authors (conducted) or based on publicly available cohort GWAS summary statistics (downloaded). The meta-analysis was performed with a sample-size weighted meta-analysis using METAL (see Methods).
OSA = Obstructive sleep apnoea; UKB = UK Biobank; MVP = Million Veteran Program; FinnGen = Finnish biobank consortium; All of Us = NIH All of Us Research Program; CLSA = Canadian Longitudinal Study on Aging; AGDS = Australian Genetics of Depression Study; GBP= Australian Genetics of Bipolar Disorder Study; AGDS & GBP(Australian Cohorts) were analysed jointly due to identical questionnaire items for OSA phenotyping; Mass General Brigham Biobank = biobank formerly known as the Partners HealthCare Biobank; ICD-10 = International Classification of Diseases, 10th Revision; SNOMED = Systematised Nomenclature of Medicine.

As a conditional analysis, we generated GWAS summary statistics for OSA after conditioning on the genetic effect of BMI (OSA_mtCOJO_BMI_) using mtCOJO. The LD score regression intercept was 1.005 (SE = 0.01), with an attenuation ratio of 0.009 (SE = 0.02). In this secondary analysis, we identified 39 independent genome-wide significant loci, of which 27 loci were within a 500kb window of SNPs identified in our primary OSA meta-analysis and 12 were novel (**Supplementary Table 3**). Manhattan and quantile–quantile plots for individual cohort and meta-analysis are available in **Supplementary Figures 1-17**.

To further refine association signals and investigate allelic heterogeneity within identified loci, we performed a conditional and joint multiple-SNP analysis using GCTA-COJO. We identified 220 conditionally independent association signals surpassing genome-wide significance, which clustered into 191 COJO-defined loci (distinct from the 147 LD-clumped discovery loci) when grouped within 500 kb windows (**Supplementary Table 4**). Of these, 17 loci harbored two or more independent association signals, and five loci contained three or more distinct signals, indicating allelic heterogeneity at select genomic regions. In total, 29 additional signals were detected within the 191 COJO-defined loci. The *FTO* locus on chromosome 16 showed two independent association signals.

To characterise if and to what extent our OSA_mtCOJO_BMI_ analyses could influence locus-level effects, we compared lead variants from the primary OSA meta-analysis with their corresponding SNP effects in OSA_mtCOJO_BMI_ and with association statistics from an independent BMI GWAS (**Supplementary Tables 1 and 5**). Across the 147 independent OSA loci, effect sizes were consistently attenuated following OSA_mtCOJO_BMI_ analyses, with most variants showing partial to substantial reductions in effect size magnitude. Genetic variants showing stronger attenuation tended to have stronger BMI associations (e.g., rs1421085 in the FTO locus), whereas several retained OSA signals displayed weak or non-significant BMI associations. The persistence of association at loci such as FTO is consistent with partial attenuation of BMI-related genetic effects, rather than their complete removal. We observed no systematic reversals in effect direction across loci. In a targeted effect size directionality screen across the 39 OSA_mtCOJO_BMI_ independent loci, effect size directions were highly concordant between OSA and OSA_mtCOJO_BMI_, with minor direction reversals in two loci (rs633715 and rs17782313), both of which showed extremely strong BMI associations.

Sensitivity analyses in Europeans are described in the **Supplementary Note**. Manhattan and quantile–quantile plots for all sensitivity analyses are available in **Supplementary Figures 18-23**.

### GWAS in African Ancestry

Next, we meta-analysed African-ancestry GWAS summary data from *All of Us* and MVP cohorts for OSA, identifying six independent genome-wide significant loci (*P* < 5 × 10^−8^) (N_cases_= 46,834; N_controls_= 149,192; **Supplementary Table 6**; **Supplementary Figures 24 and 25**). The SNP-based heritability on the liability scale for this analysis was estimated at 5.35% (SE = 0.007). The trans-ethnic genome-wide genetic correlation between OSA in the African and European cohorts was high at 0.91 (SE = 0.05, *P* = 0.08), indicating a largely shared genetic architecture (*P*-value tests for deviation from 1, thus not rejecting the hypothesis of perfect genetic sharing). Manhattan and quantile–quantile plots for individual cohort and meta-analysis are available in **Supplementary Figures 26-29**. Further details for GWAS in African ancestry are available in the **Supplementary Note**.

### GWAS in Diverse Ancestries

We meta-analysed GWAS summary statistics from European, African, Admixed American, and East Asian ancestry cohorts, incorporating ancestry-specific results from the Million Veteran Program alongside our European and African meta-analyses (N_cases_= 300,368; N_controls_= 1,450,687; **Supplementary Table 1; Supplementary Figures 30 and 31**). We identified 68 independent genome-wide significant loci (**Supplementary Table 7**), of which 9 were identical to lead variants identified in the European meta-analysis and none overlapped with the six African-ancestry lead variants. A further 45 multi-ancestry lead variants were not European lead SNPs but were present within the LD-defined European loci, mapping to 43 distinct European lead loci. Genetic correlations across ancestry-specific GWAS were estimated using Popcorn. High genetic correlations were observed between European, African, and Admixed American ancestry analyses. Full results are provided in **Supplementary Table 8.**

### Functional Annotation and Gene Prioritisation

To translate genetic loci into biological function, we applied a suite of post-GWAS analyses to the European-ancestry summary statistics. Gene discovery (**Supplementary Tables 9-12**) and pathway analysis, as well as tissue and cell-type specificity analyses (**Supplementary Tables 13-17**) are described in the **Supplementary Note**.

#### Fine-mapping of gene-level associations

To further prioritise candidate causal genes from gene discovery, pathway analysis, and tissue and cell-specificity, we performed transcriptome-wide association study (TWAS)-based fine-mapping using FOCUS across 13 GTEx v8 brain tissues. Across genomic regions showing evidence of association, we identified multiple genes with high posterior inclusion probabilities (PIP > 0.9), indicating strong support for their role in mediating genetic risk for OSA (**Supplementary Table 18**). Several genes showed particularly strong evidence (PIP ≥ 0.98), including *SCAPER*, *HSD17B6*, *ANKRD27*, *EPS15*, and *COX7A2L*. Additional candidate genes, including *TFG*, *POU1F1*, and *CEPT1*, were identified particularly within basal ganglia and cortical regions.

#### Causal gene prioritisation and spatial mapping

Summary-based Mendelian Randomisation (SMR) identified five potential causal genes in the prioritised brain tissues, including *GTF2IRD2* and *SCAPER* (*P* < 7.56×10^−07^; **Supplementary Table 19**). We also observed potential causal gene candidates of nominal statistical significance (*P* < 0.05). These findings included *AS3MT* across all 13 brain tissues; *ZSCAN31* in all tissues except the cerebellar hemisphere; *CADM2* in the nucleus accumbens, caudate nucleus, putamen, substantia nigra, cervical spinal cord, and hippocampus; and *MAPT* in the cerebellum, among others (**Supplementary Table 20**).

To further explore the neural circuitry, we leveraged spatial human cortical brain tissue data through the gsMap method. This revealed that 17 prioritised genes were robustly enriched in excitatory neurons of layers 2/3, 4, and 6, and in specific inhibitory interneurons and astrocytes. Genes such as *NPEPPS* and *ETV5* showed the strongest enrichment in layer 6, while *NRXN1* and *CACNB2* were restricted to layers 2/3 (**Supplementary Tables 21-23**). Protein–protein interaction network analysis revealed distinct functional modules involving synaptic signalling, vesicle-mediated transport, and mitochondrial energy metabolism, supporting convergent biological pathways implicated in obstructive sleep apnoea (**Figure 2**). This complements SMR findings and spatial enrichment of genes in excitatory and inhibitory neurons, together indicating that the genetic risk for OSA may converge on disruption of synaptic signalling pathways within specific cortical microcircuits.

**Figure 2.**
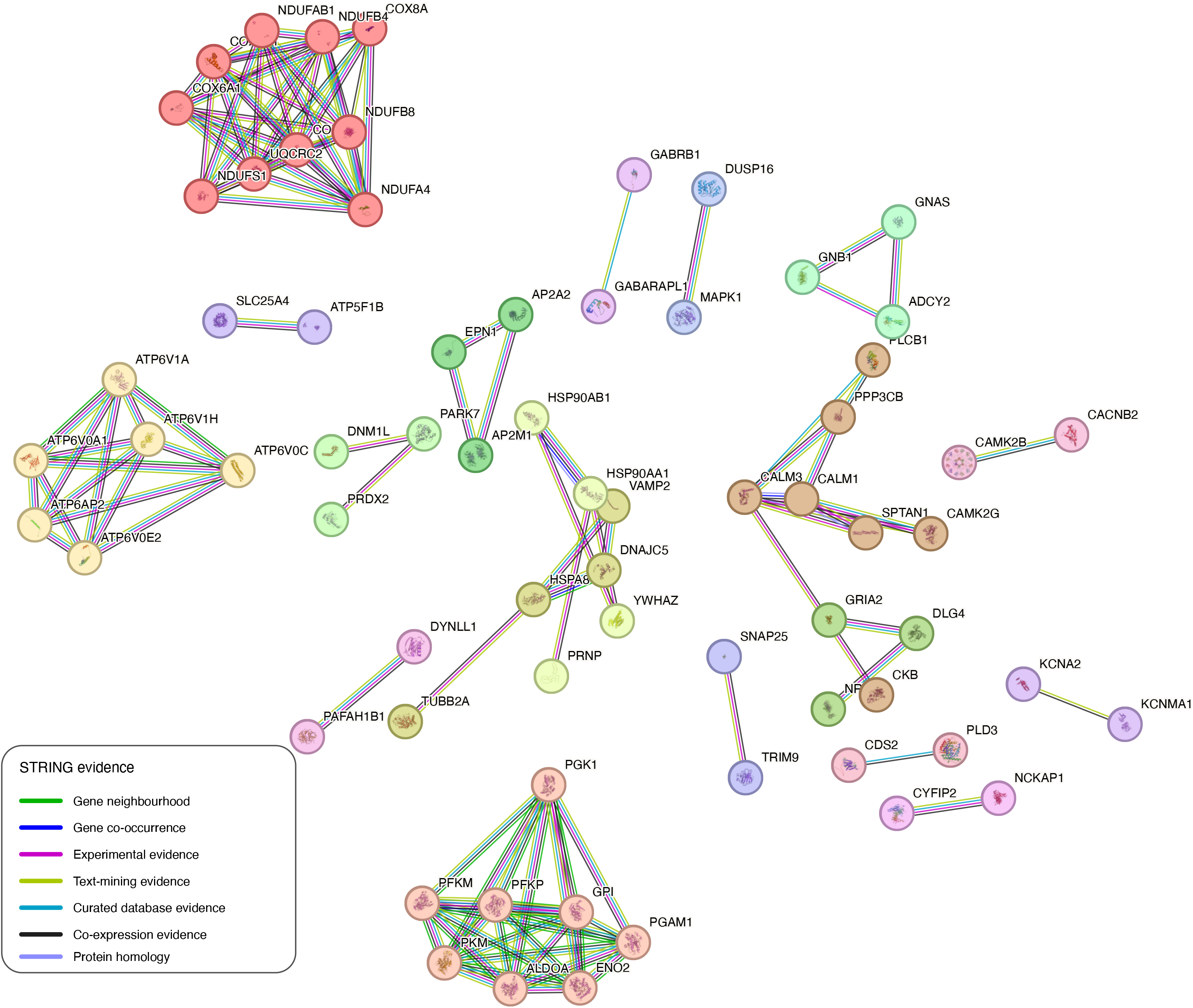
Protein–protein interaction (PPI) network of genes prioritised by gsMap in European OSA and OSA_mtCOJO_BMI_ analyses. Genes were first prioritised using gsMap based on spatial transcriptomic enrichment (Pearson correlation coefficient ≥ 0.7) in both the European OSA meta-analysis and the European OSA_mtCOJO_BMI_ analysis. Protein–protein interaction networks were then constructed using STRING v12.0. Only high-confidence interactions (STRING interaction score ≥ 0.7) are shown. Nodes represent proteins encoded by prioritised genes, and edges represent known or predicted interactions curated in STRING. Clusters of interacting proteins were identified using the Markov Cluster Algorithm (MCL) with an inflation parameter of 3. Edge colours indicate the type of evidence supporting each interaction, as defined in the key within the figure, and node colours indicate MCL clusters. The STRING output was modified for presentation by adjusting the label size and figure margins and is used under the Creative Commons Attribution 4.0 International licence (CC BY 4.0).

#### Conditional analysis removing BMI effects

As a conditional analysis, we repeated the functional annotation for the OSA_mtCOJO_BMI_ GWAS. This resulted in a marked reduction in the number of associated genes (e.g., 41 genes in MAGMA vs. 249 in the primary analysis). Notably, the significance for the well-known obesity-related gene FTO was attenuated, but not fully eliminated, and associations for *CADM2*, *ETV5*, and *AUTS2* were no longer significant after correction, consistent with partial mediation by BMI and/or residual pleiotropic effects (**Supplementary Tables 24-26**). Details for tissue-expression analyses (**Supplementary Tables 27-31**) and SMR (**Supplementary Table 32** and **33**) are available in the **Supplementary Note**.

As in the primary OSA meta-analysis, we applied gsMap to spatially resolved human cortical transcriptomic data using the OSA_mtCOJO_BMI_ GWAS. Despite the overall attenuation of association, 125 out of 637 genes showing spatial enrichment in the primary analysis remained detectable. Notably, *NRXN1, CACNB2, NPEPPS, KCNMA1, CYFIP2, TRIM9, TSPAN7, GPM6A, NDRG4, SYNE1* and *GABRB1,* continued to show spatially localised enrichment across cortical layers, particularly within excitatory neurons of layers 2/3 and 6 and select inhibitory interneuron populations (**Supplementary Tables 34 and 35**).

### Polygenic Risk Score Prediction Across Diverse Ancestries

We evaluated the predictive utility of a polygenic risk score (PRS) derived from our European GWAS in individuals of diverse ancestries (European, African, Admixed American, South Asian, East Asian, and Middle Eastern) from the *All of Us* cohort, and East Asian (Chinese) participants from the *Guangzhou Biobank Cohort Study* (GBCS) cohort.

In *All Of Us*, OSA PRS was significantly associated with clinician ascertained OSA status across all six ancestral groups analysed (N_Europeans_=220,567; N_African_=74,780; N_Admixed American_=71,178; N_Middle Eastern_=1,027; N_South Asian_=4,008; N_East Asian_=9,184; **Figure 3a**; **Supplementary Tables 36 and 37**). A clear dose-response relationship was observed; for instance, in Europeans, individuals in the top PRS decile had 2.64-fold higher odds of OSA compared to the bottom decile (**Figure 3b**). Top-decile odds were also elevated in other ancestries, with wider confidence intervals in smaller groups. Per standard deviation odds ratios (ORs) ranged from 1.14 (95% CI 1.11–1.16; *P*=2.55×10^−27^; AUC=0.65) in African ancestry to 1.47 (95% CI 1.16–1.87; *P=*1.49×10^−3^; AUC=0.72) in Middle Eastern–ancestry participants; the model including PRS, age, sex and 10 PCs had a Nagelkerke R² of up to 0.12, with similar strong effects in European (OR 1.36; *P*=2.26×10^−300^; AUC=0.65), Admixed American (OR 1.33; *P*=1.59×10^83^; AUC=0.71), East Asian (OR 1.26; *P*=2.25×10^−6^; AUC=0.70), and South Asian (OR 1.21; *P*=6.95×10^3^; AUC=0.75) groups. These associations were attenuated but remained largely significant after conditioning on BMI. The PRS also correlated with objective, FitBit-derived sleep metrics (e.g., fragmentation index, number of awakenings, REM sleep percentage, sleep efficiency, and wake after sleep onset; *see Methods* and **Supplementary Table 38**). In Europeans (N_Europeans_=8,779), OSA PRS correlated with more frequent awakenings (*P* = 2.46 × 10^−4^) and wake after sleep onset (WASO) (*P* = 2.87 × 10^−2^). Associations for other ancestry groups (N_Admixed American_=771; N_African_=617; N_East Asian_=340; N_South Asian_=112; N_Middle Eastern_=27) were largely non-significant (*P* > 0.05) yet showed similar r-squared values.

**Figure 3.**
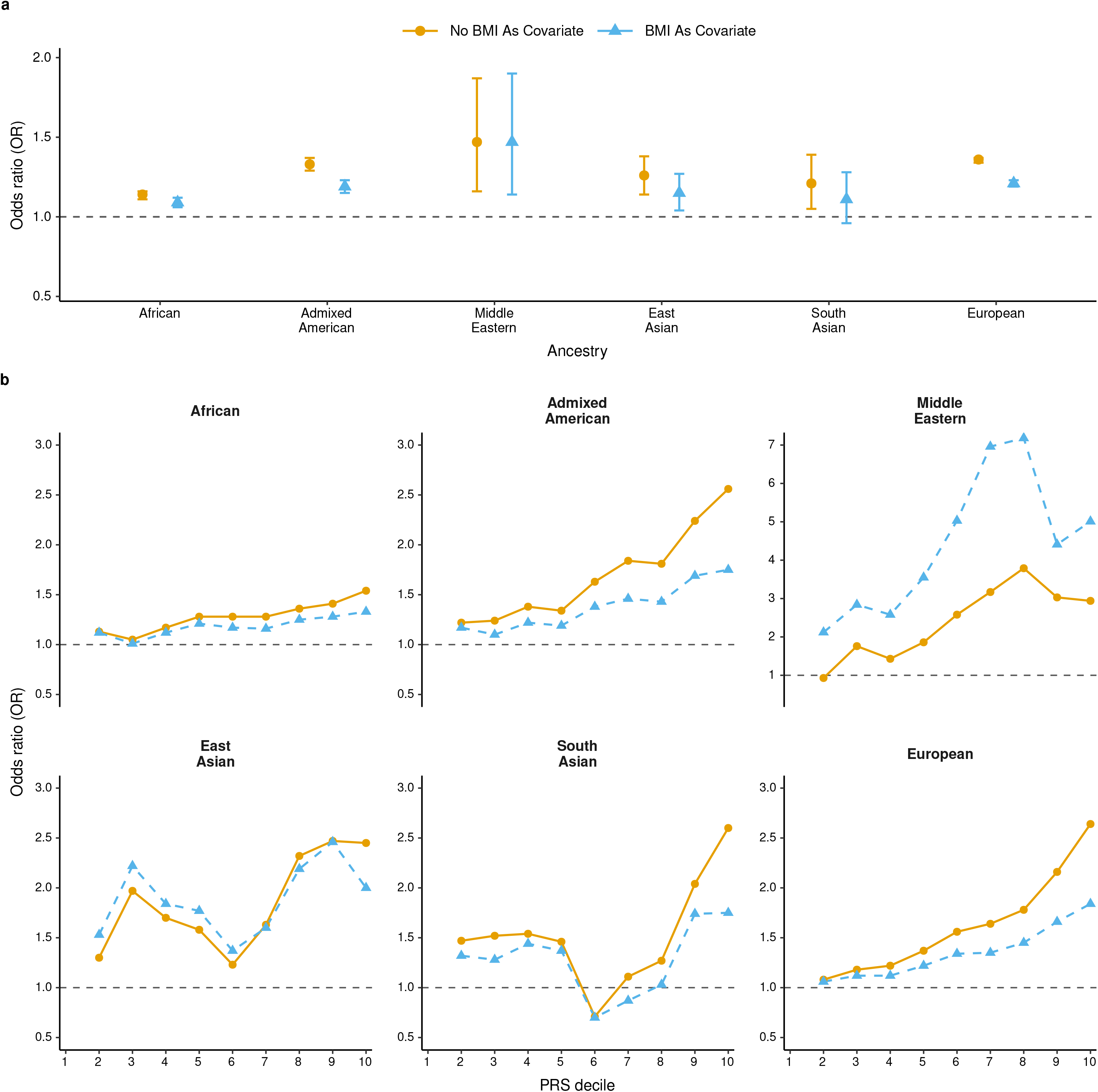
Polygenic prediction in populations from diverse ancestral backgrounds in the All Of Us cohort. Clinician ascertained obstructive sleep apnoea (OSA) status polygenic risk scores (PRS) were derived using *SBayesRC.* Odds ratios (ORs) were estimated with logistic regressions. **a)** Per standard deviation ORs for clinician ascertained OSA in each ancestry, including European (N = 220,567), African (N = 74,780), Admixed American (N = 71,178), East Asian (N = 9,184), South Asian (N = 4,008), and Middle Eastern (N = 1,027). The corresponding sample sizes with BMI adjustment were 188,005, 72,066, 64,360, 8,250, 3,592 and 949, respectively. Markers represent odds-ratio estimates per standard-deviation increase in the OSA PRS from logistic regression, and error bars indicate 95% confidence intervals. The horizontal dashed line indicates the null value (OR = 1). **b)** Risk stratification by PRS decile: ORs in deciles 2–10 (vs. decile 1) for each ancestry. In both a) and b), orange circles show models without body mass index (BMI) adjustment; blue triangles show models with BMI as a covariate.

In the East Asian GBCS cohort, each standard deviation increase in the European derived OSA PRS was associated with a 10% higher odds of having a phenotypic OSA risk score ≥ 2 on the 4-item STOP questionnaire (*see Methods*; N_TOTAL_ = 2,023; 863 cases; OR = 1.10; 95% CI 1.0–1.2; *P* = 0.01; R² = 0.002), independent of age and sex. This effect persisted (OR = 1.10; *P* = 0.01; R² = 0.002) after further adjustment for BMI. Similar results were observed when defining OSA risk with a STOP score ≥ 3 (526 cases; OR = 1.10; 95% CI 1.0–1.2; *P* = 0.01; R² = 0.002), both before and after BMI adjustment. For self-reported snoring (N_TOTAL_ = 2,023; 891 cases), OSA PRS conferred a 10% increase in odds per standard deviation (OR = 1.10; 95% CI 1.0–1.2; *P* = 0.008; R² ≈ 0.003), even after including BMI as a covariate in the model (**Supplementary Table 39**).

### Genetic Overlap and Causality with Other Complex Traits

**Genome-wide genetic correlations.** We estimated the genetic correlation (*r*_G_) between OSA and 114 complex traits (**Figure 4; Supplementary Table 40**). After Bonferroni correction (*P* < 2.19 x 10^−04^; *see Methods*), OSA showed significant positive genetic correlations with neuropsychiatric disorders (attention-deficit/hyperactivity disorder (ADHD), anxiety, major depressive disorder) cardiometabolic traits (BMI, stroke, type 2 diabetes), and other sleep-related phenotypes (snoring, napping, long/short sleep duration). We also observed significant negative correlations (*P* < 2.19 x 10^−04^) with anorexia nervosa, schizophrenia, and HDL cholesterol. In addition, we estimated genetic correlations among the individual OSA cohorts included in the meta-analysis, which revealed predominantly positive and moderate-to-high genetic correlations, supporting a shared genetic architecture across cohorts (**Supplementary Figure 32; Supplementary Table 41**).

**Figure 4.**
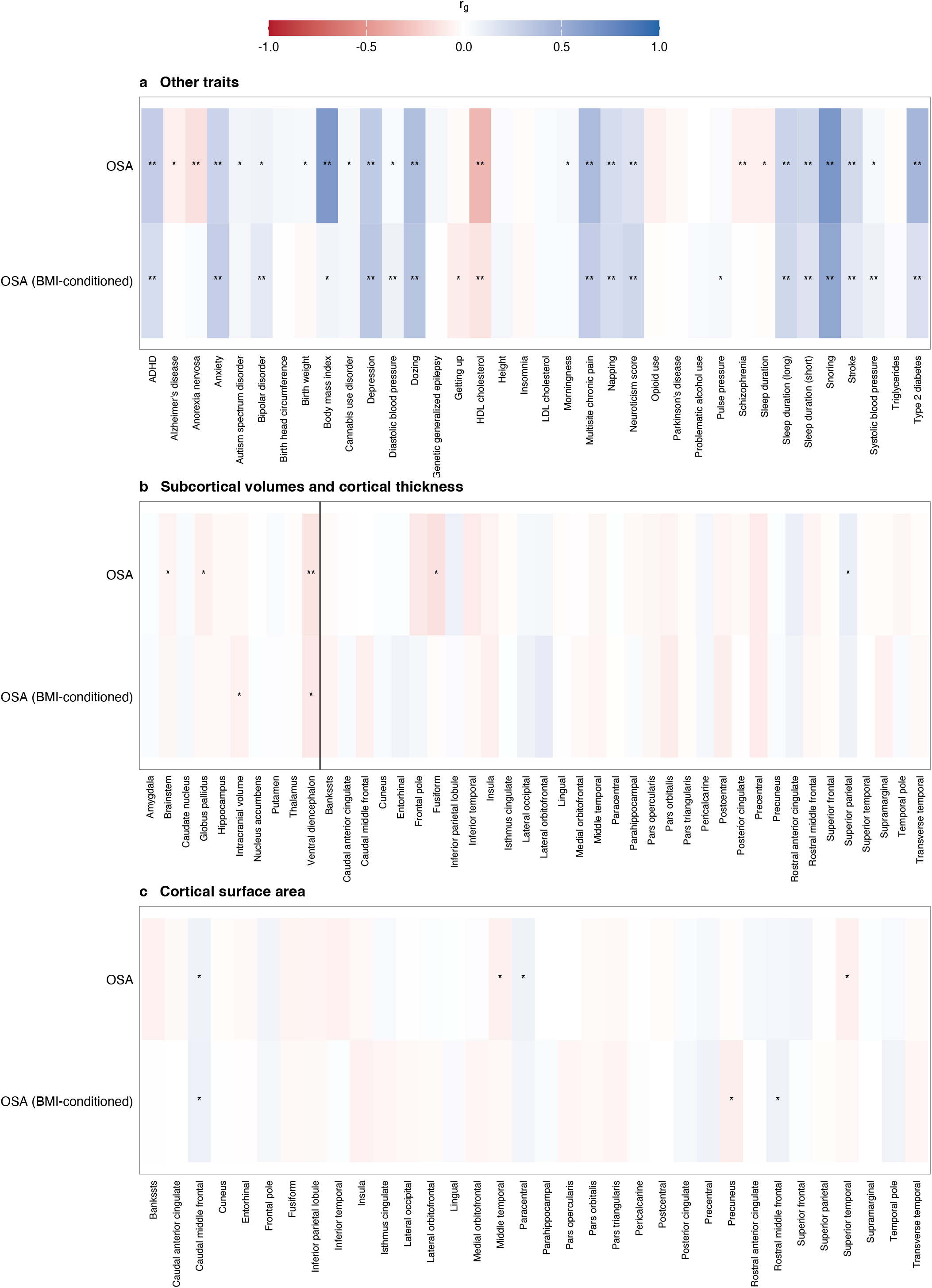
Genetic overlap with other complex human traits. Heatmaps depict genetic correlations (rG) of obstructive sleep apnoea (OSA) and OSA conditioned on body mass index using mtCOJO (OSA (BMI-conditioned)) with **a)**, other complex traits; **b)**, subcortical volumes and cortical thickness; and **c)**, cortical surface area. In **b)**, the vertical line separates subcortical volumes from cortical thickness measures. Genetic correlations were estimated using LD score regression. P values were calculated using two-sided chi-squared tests with one degree of freedom. *P < 0.05; **P < 2.19 × 10−4, corresponding to Bonferroni correction for 228 genetic-correlation tests. Exact P values and standard errors are provided in Supplementary Table 40.

As a conditional analysis, we investigated if and to what extent removing the effect of BMI influenced the genetic correlations of OSA with complex human traits. The genetic correlation between OSA and OSA_mtCOJO_BMI_ was 0.82 (SE = 0.01, *P* < 1 x 10^−300^) while the estimate for OSA_mtCOJO_BMI_ and BMI was 0.06 (SE = 0.02, P < 6.28 x 10^−04^). OSA_mtCOJO_BMI_ results are, for the most part, consistent with OSA’s, with slight reductions in the magnitude of the genetic correlations. When compared to OSA results, although most of the following genetic correlations maintained the same direction albeit a reduction in their magnitude, we observed the absence of nominal genetic correlations (*P* < 0.05) with autism spectrum disorder, Alzheimer’s disease, birthweight, cannabis use disorder, subcortical brainstem and globus pallidus volumes, and some brain cortical measurements. Similarly, genetic correlations with anorexia nervosa and schizophrenia were no longer statistically significant after multiple testing corrections.

Colocalisation analyses (**Supplementary Table 42**), are described in the **Supplementary Note**.

#### Mendelian randomisation

To investigate potential causal relationships, we used Latent Heritable Confounder Mendelian Randomisation (LHC-MR). After multiple testing corrections (*P* < 1.47 X 10^−03^; **Supplementary Table 43**), we found evidence that genetic liability for ADHD and depression increased the risk for OSA. Conversely, genetic liability for OSA appeared to increase the risk for multisite chronic pain, type 2 diabetes, and snoring.

We also observed bidirectional relationships between OSA and daytime dozing, napping, HDL cholesterol, schizophrenia, and, notably, with BMI, where the influence of BMI on OSA risk showed stronger statistical evidence (i.e., smaller *P*) yet smaller effect magnitude than the reverse.

## DISCUSSION

In the largest GWAS meta-analysis of obstructive sleep apnoea (OSA) to date, including over 1.4 million individuals from 5 countries (Australia, USA, UK, Canada, and Finland), we identified 147 independent genome-wide significant loci (P < 5 × 10^−8^), of which 141 had not been reported previously^6,7^. LD score regression intercepts suggest that the elevated lambda and inflation in the quantile plots are most likely due to polygenicity rather than population stratification. Similarly, attenuation ratios indicate correct genomic control. Our SNP-based heritability estimate of 16% is substantially higher than previous reports (8-13%)^6,7^. Crucially, our OSA_mtCOJO_BMI_ analyses, which identified 39 independent genome-wide significant loci, provide substantial insights into the genetic basis of OSA beyond the well-established effects of BMI. Through in-depth functional annotation, we prioritised genes and implicated specific biological pathways and tissues. We demonstrated that a European-derived polygenic risk score (PRS) for OSA is robustly associated with clinician ascertained OSA status, Fitbit sleep features, and symptom-based OSA risk and snoring assessed via the 4-item STOP questionnaire, with its predictive power varying across ancestries. We also clarified the genetic relationships between OSA and a range of complex health traits. Given the well-established role of adiposity in the aetiology of OSA, in this section we place emphasis on results from the OSA_mtCOJO_BMI_ GWAS, particularly when discussing functional annotation and gene prioritisation results, to distinguish genetic effects mediated through obesity from those potentially acting through alternative biological pathways.

Fine-grained spatial gene analyses revealed layer and cell type–specific expression patterns in the human cortex in both the primary and the OSA_mtCOJO_BMI_ analyses, with signals concentrated in layer 6 excitatory neurons, and other signals in layers 2/3 and 4, inhibitory interneuron subtypes (Inh.1–3), and astrocyte subtype 2. We highlight that Layer 6 is a key regulator of corticothalamic feedback, which governs slow oscillations and sleep spindles essential for sleep-stage regulation and memory consolidation^12,13^. Notably, several genes showed robust replication across analyses, including *NPEPPS, KCNMA1, CYFIP2, TRIM9, TSPAN7, GPM6A, NDRG4,* and *SYNE1*, with enrichment primarily in deep cortical layers and excitatory neuronal populations. The products of these genes are involved in synaptic maintenance, cytoskeletal dynamics, neuronal excitability, and axonal organisation, processes that may influence cortical control of arousal and respiratory stability independent of adiposity. The persistence of these spatial signals despite BMI adjustment suggests that aspects of cortical microcircuitry are likely to contribute to OSA susceptibility beyond structural airway or metabolic mechanisms. Discussion for gene-level and tissue-specific analyses is available in the **Supplementary Note**.

Among the replicated genes, several have well-established roles in synaptic transmission and sleep-relevant neural function. *NRXN1*, encodes neurexin-1, a synaptic adhesion protein involved in excitatory synapse specification and neurotransmitter release^14^, while *CACNB2* encodes the beta-2 auxiliary subunit of voltage-gated calcium channels, which regulates channel activity and presynaptic calcium influx^15^, both critical for synaptic signalling and network excitability. In addition, *KCNMA1* encodes a large-conductance calcium-activated potassium channel, modulating neuronal firing and has been implicated in sleep–wake regulation and respiratory control^16^. Additional genes such as *GABRB1*, expressed in astrocytic and inhibitory neuronal contexts, further point to a role for excitatory–inhibitory balance and neuromodulatory support in shaping sleep stability^17^. Our findings suggest that genetic susceptibility for OSA, even after conditioning on BMI, implicates components of cortical and corticothalamic circuitry involved in synaptic signalling and neuronal excitability rather than broad tissue-wide effects. In particular, the convergence on genes regulating ion channel activity, inhibitory neurotransmission, and synaptic maintenance is consistent with altered excitatory–inhibitory balance and network stability in pathways governing arousal regulation and sleep continuity. While these effects are more circumscribed than those observed in the primary analysis, they point to biologically plausible neural mechanisms that may contribute to sleep fragmentation and impaired restorative sleep in OSA beyond adiposity-related pathways. These observations provide testable hypotheses for future functional investigations using animal or cellular models.

We demonstrated that European-derived polygenic risk scores for OSA predict clinician ascertained OSA status, some Fitbit-derived sleep features, and symptom-based OSA risk and snoring assessed via the 4-item STOP questionnaire, even when including BMI as a covariate in the predictive models. We show that each SD increase in PRS confers 14 - 47% higher odds of clinically ascertained OSA in European, African, Admixed American, South Asian, East Asian, and Middle Eastern participants. When stratifying each ancestry specific PRS distribution into deciles, we observed that individuals in the top PRS decile had approximately 1.5–3-fold higher odds of OSA when compared to the bottom decile. Moreover, PRS correlations with continuous Fitbit metrics were largely non-statistically significant for non-Europeans. This finding is most likely explained by the substantial differences in sample sizes across ancestries, as only a small fraction of the All of Us participants have Fitbit data available (*see Methods*). Related to this, in the independent Chinese cohort, OSA PRS modestly predicted STOP questionnaire OSA risk scores and self reported snoring, confirming that European derived PRS retained predictive power across ancestries.

Previous studies have aimed to investigate the genetic overlap of OSA with complex human traits^6,7^. In the present study, we identified genetic correlations between OSA and 17 complex human traits, including psychiatric, anthropometric, cardiometabolic, and other sleep-related phenotypes. Further interrogation of these associations uncovered potential causal genetic effects in which genetic liability to ADHD and depression tends to increase OSA risk, mirroring clinical observations where these disorders often co occur and may share neurobiological pathways^18,19^. Conversely, genetic susceptibility to OSA may contribute to the development of cardiometabolic and pain-related phenotypes, elevating the risk for multisite chronic pain, type 2 diabetes, and snoring, which is consistent with systemic effects of intermittent hypoxia and sleep fragmentation on inflammation^20,21^, reduced insulin sensitivity^22^, and autonomic regulation^23^. Our bidirectional LHC MR results with BMI underscore a self reinforcing obesity–OSA cycle, in which adiposity mechanically promotes airway collapse while OSA could reciprocally worsen energy balance via hormonal dysregulation^24^. The relationships between OSA and HDL cholesterol point to lipid metabolism as both a contributor and a consequence of OSA pathophysiology. Similarly, findings of nominal statistical significance for anxiety, stroke, ventral diencephalon volume, and anorexia nervosa could provide hints at additional neurovascular and neurodevelopmental axes influencing OSA susceptibility that warrant further investigation. Our observations highlight the relevance of comorbid psychiatric conditions in the aetiology of OSA, which may inform screening, diagnosis and management.

It is well-established that increased adiposity is a risk factor for OSA, partly through mechanisms such as upper airway fat deposition and reduced lung volume.^25^ However, the genetic architecture of OSA is not solely driven by BMI-related effects^6,7^, and while adipokine signalling has been proposed as a link,^26,27^ associations with related genes (e.g., *LEP*, *ADIPOQ*, *RETN*) were not prominent in our results. Our analysis conditioning on BMI (i.e., OSA_mtCOJO_BMI_) suggests that OSA has a substantial genetic component that is not fully explained by BMI-related genetic effects, while also retaining residual BMI-associated signal at specific loci, consistent with the established role of adiposity in OSA pathophysiology.

As expected, this analysis reduced the number of significant loci from 147 to 39, and attenuated the signal for key metabolic genes, including *FTO*. Crucially, this still represents a major advance over previous BMI-conditioned analyses, which identified far fewer loci. Most importantly, while metabolic signals were reduced, biological pathways implicating neurogenesis related processes persisted, and eQTL mapping continued to implicate the same brain structures (albeit with fewer and more specific candidate genes). Furthermore, and in contrast to previous studies^7^, the genetic correlations profile for OSA_mtCOJO_BMI_ largely mirrored our primary results. Taken together, these findings provide compelling evidence that while BMI is a major contributor to OSA aetiology, a distinct and prominent neurobiological architecture independently contributes to individual risk. We also note that the persistence of attenuated signals at loci such as *FTO* should be interpreted cautiously, as BMI is an imperfect proxy for adiposity and OSA_mtCOJO_BMI_ analyses at the summary-statistics level cannot fully disentangle pleiotropy from residual confounding.

Regarding conditioning on BMI in the present study, we note that mtCOJO conditions on the genetic component of BMI using GWAS summary statistics rather than adjusting for measured BMI at the individual-participant level^28^. This approach aims to attenuate genetic effects shared between BMI and OSA while reducing bias that may arise when adjusting for a heritable and partially mediating trait, such as BMI, in individual-level GWAS models. Specifically, conditioning on measured BMI can induce collider bias by distorting associations between genetic variants and OSA when those variants influence BMI and other related metabolic traits.

Consistent with our mtCOJO approach, OSA_mtCOJO_BMI_ retained stable genome-wide association signals, including preserved effect-size estimates and genetic signals outside major metabolic-related regions, while substantially reducing shared genetic signal with BMI. LD score regression applied to OSA_mtCOJO_BMI_ summary statistics showed an intercept close to one and a low attenuation ratio, indicating no evidence of inflation or additional confounding introduced by the conditioning approach. Therefore, our results support mtCOJO as a valid approach for downstream analyses, including genetic correlation and causal inference, when interpreted as reflecting substantial attenuation of BMI-shared genetic effects rather than full BMI-independent associations. The limitations of the present study are discussed in the **Supplementary Note**.

In conclusion, this study fundamentally refines our understanding of the genetic architecture of OSA, demonstrating a complex polygenic basis in which adiposity plays a prominent role while a subset of genetic risk extends beyond BMI to implicate neurobiological pathways involved in synaptic function and corticothalamic circuitry. The predictive performance of our polygenic risk scores across diverse ancestries highlights its potential to capture genetic liability for OSA. However, its modest discriminative ability indicates that it is not yet suitable as a standalone clinical prediction tool and should be interpreted in the context of established risk factors, such as BMI. Nevertheless, these findings represent an important step towards integrating genetic information into OSA research and risk modelling. Critically, our findings provide a rich atlas of testable hypotheses for the field; future research must now focus on functionally validating these novel genes and pathways to mechanistically dissect their roles in sleep fragmentation and arousal. Expanding discovery efforts in non-European populations will be paramount to ensuring that the benefits of genomic discovery are equitable. Ultimately, this work paves the way for future therapeutic strategies aimed at the molecular roots of OSA, offering hope for the millions affected by this debilitating condition.

## ACKNOWLEDGEMENTS

This research used data from UK Biobank under application number 25331. We acknowledge the Mass General Brigham Biobank for providing samples, genomic data and health information. We gratefully acknowledge All of Us participants for their contributions, without whom this research would not have been possible. We also thank the National Institutes of Health’s All of Us Research Program for making available the participant data examined in this study.

We acknowledge the participants and investigators of the FinnGen study. We acknowledge the Million Veteran Program (MVP), Office of Research and Development, Veterans Health Administration, and its participants, and thank the MVP investigators for making GWAS summary statistics available; this publication does not represent the views of the Department of Veterans Affairs or the United States Government. This research used data and biospecimens collected by the Canadian Longitudinal Study on Aging (CLSA), including the Baseline Comprehensive Dataset version 4.0, Follow-up 1 Comprehensive Dataset version 1.0 and Genetic Data version 3.0, under application number 190225. The CLSA is led by Parminder Raina, Christina Wolfson and Susan Kirkland. We gratefully acknowledge the time and commitment of the CLSA participants, without whom this research would not have been possible. The opinions expressed in this manuscript are those of the authors and do not reflect the views of the CLSA or any affiliated institution.

We thank all investigators of the Guangzhou Biobank Cohort Study for their generous contribution of genetic and longitudinal data. We thank the participants of the Genetics of Bipolar Disorder project for their time and support. We also thank all participants who contributed to the Australian Genetics of Depression Study.

We acknowledge everyone who assisted with the conception, implementation, beta testing, media campaign and data cleaning for the Australian Genetics of Depression Study. In particular, we thank Ken Kendler, Patrick Sullivan, Andrew McIntosh and Cathryn Lewis for their input on the questionnaire; Lorelle Nunn, Mary Ferguson, Lucy Winkler and Natalie Garden for data and sample collection; Natalia Zmicerevska, Alissa Nichles and Candace Brennan for participant recruitment support; Jonathan Davies, Luke Lowrey and Valeriano Antonini for support with information technology; and Vera Morgan and Ken Kirkby for assistance with the media campaign.

## FUNDING

The Canadian Longitudinal Study on Aging is funded by the Government of Canada through the Canadian Institutes of Health Research (grant LSA 94473) and the Canada Foundation for Innovation, as well as by the provinces of Newfoundland and Labrador, Nova Scotia, Quebec, Ontario, Manitoba, Alberta and British Columbia.

Data collection for the Australian Genetics of Depression Study was supported by the Australian National Health and Medical Research Council (NHMRC) through grant GNT1086683 awarded to N.G.M.

V.F.O. acknowledges PhD scholarship support from the Global Parkinson’s Genetics Program (GP2). B.U. is supported by a Higher Degree Research Tuition Fee Scholarship from Queensland University of Technology and a scholarship from the QIMR Computational Neurogenomics Laboratory. F.C. is supported by scholarships from the University of Queensland and the QIMR Computational Neurogenomics Laboratory. E.M.B. is supported by the University of Queensland Health Research Accelerator Program. D.S. is supported by a University of Queensland Amplify Professorial Research Fellowship. I.B.H. is supported by the research fellowship “Right care, first time: delivering technology-enabled mental health care to young people at scale” (2023–2027; APP2016346). M.E.R. acknowledges support from the Rebecca L. Cooper Medical Research Foundation (F20231230). S.M. is supported by an Australian NHMRC Investigator Grant (2034568).

Data collection for the Genetics of Bipolar Disorder project was funded, and data analysis was supported, by the Australian NHMRC through grant APP1138514 awarded to S.E.M. S.E.M. is also supported by an NHMRC Investigator Grant (APP2025674).

The funders had no role in study design, data collection and analysis, decision to publish or preparation of the manuscript.

## AUTHOR CONTRIBUTIONS

**Core analysis and writing group:** LMGM, ZC, SDT, AP, JWX, VFO, AMA, BU, MMM, XH, CP, AAR, FC, NSO, MS, MER

**Data collection:** DJS, PAL, SEM, IBH, EMB, NGM, SM, XD, SJL, MS, JW, MER

**Contributed to the editing of the manuscript:** LMGM, ZC, SDT, AP, JWX, VFO, AMA, BU, MMM, XH, CP, AAR, FC, NSO, DJS, PAL, SEM, IBH, EMB, NGM, SM, XD, SJL, MS, JW, MER

## COMPETING INTERESTS

IBH holds a 3.2% equity interest in Innowell Pty Ltd, a company focused on the digital transformation of mental health services. DJS serves on the Viatris Australian Clozapine Quality Advisory Committee and has received speaker and/or consultancy fees from Servier, Otsuka, Viatris and Lundbeck. These entities had no role in the design, conduct, analysis, interpretation or reporting of this study. The other authors declare no competing interests.

## METHODS

### Ethics Statement

This research complied with all relevant ethical regulations. All contributing cohorts obtained approval from their respective institutional review boards or ethics committees, and all participants provided informed consent. The analyses of data from the Australian Genetics of Bipolar Disorder Study and the Australian Genetics of Depression Study were approved by the QIMR Berghofer Medical Research Institute Human Research Ethics Committee under protocols P3408 and P2118, respectively. UK Biobank received ethical approval from the North West Multi-centre Research Ethics Committee (reference 16/NW/0274), and this research was conducted under UK Biobank application 25331. The All of Us Research Program protocol was reviewed by the All of Us Research Program Institutional Review Board, and analyses were conducted on the Controlled Tier Dataset version 8 in accordance with the program’s data use agreement. Canadian Longitudinal Study on Aging data were used under application 190225 in accordance with the CLSA data access agreement. The Guangzhou Biobank Cohort Study was approved by the Guangzhou Medical Ethics Committee of the Chinese Medical Association, and all participants gave written informed consent. The remaining analyses used previously collected controlled-access or publicly available data and GWAS summary statistics under the relevant cohort-specific ethical approvals and data-use agreements.

### Statistics

In this study we performed several statistical approaches, including linear regression, linear mixed-effects associations, genome-wide association studies, LD-score regression, and METAL-based meta-analysis of GWAS summary statistics. Each approach is described in detail below. GWAS analyses for individual cohorts in the present study were conducted with either SAIGE(v1.3.0)^29^ or REGENIE (v4.1.2; v2.2.4 for the CLSA GWAS and v3.2.6 for the All of Us African-ancestry GWAS)^30^. We used the same parameters and quality control procedures as the GWAS summary data that were leveraged from public repositories (e.g. FinnGen and Million Veteran Program) to ensure consistency. Details for each individual cohort and the meta-analyses are provided below.

### Cohorts and GWAS for European populations

Details for cohorts (All of Us, Australian cohorts, Canadian Longitudinal Study on Aging, UK Biobank, FinnGen, Million Veteran Program, and Mass General Brigham Biobank) and individual GWAS for European populations are provided in the **Supplementary Note**.

### GWAS meta-analysis in European populations

#### METAL

We used METAL^31^ (v20200505) to perform a sample-size weighted (p-value based) meta-analysis for OSA using data from the cohorts described previously (N_cases_= 232,292; N_controls_= 1,259,390; N_total_= 1,491,682; **Table 1**). All analyses were restricted to autosomal variants. Following the well-established recommendations for case-control imbalance, cohorts were weighted according to their effective sample size as described by the equation: Neff = 4/(1/Ncases + 1/Ncontrols)^31^.

All contributing GWAS underwent cohort-level quality control prior to meta-analysis, including restriction to autosomal variants, exclusion of variants with minor allele frequency (MAF) < 0.01 and imputation quality (INFO) < 0.60, and adjustment for sex, age, genotyping array (where applicable), and the first 10 principal components of ancestry. Summary statistics were harmonised to a common set of fields (SNP, A1, A2, BETA, SE, P, and allele frequency), and genomic positions are reported on the GRCh37 (hg19) build. In METAL, the flags AVERAGEFREQ and MINMAXFREQ were enabled to compute average and minimum/maximum allele frequencies across cohorts, facilitating detection of variants with substantial allele-frequency discrepancies. Variants failing cohort-level MAF thresholds were excluded prior to meta-analysis, and a consistent MAF threshold of 0.01 was additionally applied during post-processing of the meta-analysis results. Between-cohort heterogeneity was assessed using Cochran’s Q statistic as implemented in METAL. For each variant, heterogeneity metrics including the Q statistic, I², and heterogeneity P-value were computed and are reported in **Supplementary Table 1**. Details for genomic control, LD score regression intercept, and attenuation ratio are available in the **Supplementary Note**.

Independent loci for OSA were determined by performing a conservative clumping procedure in PLINK 1.9^32^ (*P_*1 = 1 × 10^−8^, *P_*2 = 1 × 10^−5^, *r*2 = 1 × 10^−3^, kb = 1,000). Independent genome-wide loci not reported in previous studies are claimed based on a comparison of the independent unique loci identified in the present study against independent genome-wide significant loci for OSA^6,7^ reported in previous studies based on exact rs numbers. We considered LD information in the definition of the independent genome-wide loci not reported in previous studies by performing a clumping procedure using PLINK 1.9^32^ (*P_*1 = 1 × 10^−8^, *P_*2 = 1 × 10^−5^, *r*2 = 1 × 10^−3^, kb = 1,000).

#### Conditional and Joint Analysis (GCTA-COJO)

To identify loci harboring multiple independent association signals, we performed conditional and joint multiple-SNP analysis using GCTA-COJO(v1.94.1)^33^. Analyses were conducted separately for each chromosome using genome-wide significant variants (*P* < 5 × 10^−08^), applying a stepwise model selection procedure with a 2 Mb window and a collinearity threshold of 0.9. Variants with minor allele frequency <1% were excluded. Joint effect estimates and corresponding conditional p-values were derived after adjusting for other selected variants within each region.

Independent signals were defined as variants with conditional *P* < 5 × 10^−08^. To summarise locus-level architecture, conditionally independent variants were clustered within ±500 kb windows. The FTO locus was defined as the genomic interval spanning chromosome 16: 53,737,875–54,155,853 base pairs^34^ for locus-specific evaluation of allelic heterogeneity.

#### Sensitivity analyses

All details to assess the robustness of the meta-analysis to potential phenotype heterogeneity and cohort-specific effects are available in the **Supplementary Note**.

### Conditional analysis

#### Body mass index genetic effect attenuation

Body mass index (BMI) is an established risk factor for OSA, and genetic variants that influence BMI are also likely to contribute to OSA risk. However, previous studies have suggested that genetic factors may influence the aetiology of OSA independently of BMI^6–8^. In the present study, we sought to further interrogate the effect of BMI on OSA. Therefore, we leveraged a set of publicly available GWAS summary statistics for BMI, including ∼1.1 million participants^35^ and multi-trait-based conditional and joint analysis (mtCOJO)^28^ using GCTA (v1.91.7), to adjust for BMI while avoiding biases due to collider bias (i.e., the emergence of a spurious association between a pair of variables when a common outcome is modeled as a covariate). With this approach, we aimed to estimate, for each SNP, the association with OSA that was independent of the SNP’s association with BMI. In addition, to estimate independent genome-wide loci that were common for both the OSA_mtCOJO_BMI_ and our original OSA meta-analyses, we compared SNPs identified via clumping (performed as described above) in both datasets. Specifically, common independent genome-wide loci were defined either by exact rsID match or by genomic proximity, where a SNP from OSA_mtCOJO_BMI_ was considered common with the original OSA meta-analysis if it fell within ±500 kb of any independent genome-wide SNP from the original OSA analysis. We note that mtCOJO operates at the summary-statistics level and estimates SNP effects conditional on the genetic component of BMI, rather than providing fully BMI-adjusted associations derived from individual-level data. As such, this approach does not account for environmental pathways influencing BMI and should not be interpreted as reflecting complete BMI-independent effects. Details on how we evaluated the locus-level behaviour of BMI conditioning and assess potential collider-like artefacts is available in the **Supplementary Note**.

### Cohorts and GWAS for African populations

Details for cohorts (All of Us and Million Veteran Program) and individual GWAS for African populations are provided in the **Supplementary Note**.

### GWAS meta-analysis in African populations

#### METAL

We used METAL^31^ (v20200505) to perform a sample-size weighted (p-value based) meta-analysis for OSA using data from the cohorts described previously (N_cases_= 46,834; N_controls_= 149,192; N_total_= 196,026). Following the well-established recommendations for case-control imbalance, cohorts were weighted according to their effective sample size as described by the equation: Neff = 4/(1/Ncases + 1/Ncontrols)^31^. Independent loci for OSA were determined by performing a conservative clumping procedure in PLINK 1.9^32^ (*P_*1 = 5 × 10^−8^, *P_*2 = 1 × 10^−5^, *r*2 = 0.05, kb = 1,000). For each variant, heterogeneity metrics including the Q statistic, I², and heterogeneity P-value were computed and are reported in **Supplementary Table 1**.

### Transethnic GWAS meta-analysis

We conducted a transethnic GWAS meta-analysis for OSA by combining ancestry-specific GWAS summary statistics from European, African, Admixed American, and East Asian populations. This analysis integrated the European and African ancestry meta-analyses described above together with ancestry-stratified GWAS results from the Million Veteran Program (MVP) for Admixed American and East Asian participants (**Supplementary Table 44**). Meta-analysis was performed using METAL^31^ (v20200505) with a sample-size weighted (p-value based) approach, with each cohort weighted by its effective sample size to account for case–control imbalance, calculated as Neff = 4/(1/Ncases + 1/Ncontrols). All contributing GWAS summary statistics underwent the same quality control, harmonisation, and meta-analysis procedures as described for the European and African ancestry GWAS analyses, including restriction to autosomal variants, application of standard variant-level filters, and harmonisation of effect estimates and allele alignment. Independent genome-wide significant loci were identified using a conservative LD-based clumping procedure in PLINK 1.9^32^ (P_1 = 1 × 10−8, P_2 = 1 × 10−5, r2 = 0.05, kb = 1,000), applied to the transethnic meta-analysis results using a multi-ancestry 1000 Genomes reference panel (∼2,500 individuals). Genome-wide significant loci identified in the multi-ancestry analysis were compared against ancestry-specific results to assess locus sharing and potential ancestry-specific signals. For each variant, heterogeneity metrics including the Q statistic, I², and heterogeneity P-value were computed and are reported in **Supplementary Table 1**.

### Transethnic genetic correlation

#### Popcorn

We leveraged GWAS summary statistics for OSA in individuals of European and African ancestries generated in the present study, as well as Admixed American and East Asian ancestry GWAS summary statistics from the Million Veteran Program, to estimate the transethnic genetic correlation using Popcorn(v1.0)^36^. Popcorn models the covariance of SNP effect sizes across two populations by regressing the product of Z scores on cross-population LD scores, adjusting for differences in linkage disequilibrium and allele frequency. By comparing this covariance to the heritability within each ancestry, we derived a trans ethnic genetic correlation that quantifies the proportion of shared genetic architecture between the traits in GWAS with different ancestries^36^. The p-value reported by Popcorn tests whether the genetic correlation is significantly less than 1. A large p-value (e.g., >= 0.05) implies not rejecting the hypothesis of perfect genetic sharing between populations^36^.

### Functional annotation and gene prioritisation

We performed functional annotation and gene prioritisation analyses. Specifically, these analyses included MAGMA, fastBAT, LDSC-SEG, eQTL mapping, FOCUS, Summary-based Mendelian Randomisation (SMR), which are described in detail in the **Supplementary Note**, and spatial mapping of cells in cortical brain tissue (gsMap).

Considering our gene discovery and tissue enrichment findings, we further investigated the genes prioritised in the gene discovery phase via spatial transcriptomics analyses using gsMap (v1.71.2)^37^. We leveraged human cortical brain tissues to investigate fine-grained spatial gene expression patterns underlying OSA’s neural circuitry based on the results from our tissue expression analyses described above. We filtered our results for those involving genes that were identified in MAGMA or fastBAT analyses. We implemented a Bonferroni multiple-testing comparison based on the number of tested cell types (0.05 / 14 = 3.57×10^−03^), and a strict Pearson correlation coefficient (PCC) between the modelled expression and true expression (PCC > 0.70).

As a conditional post-GWAS analysis, to further understand the influence of BMI in OSA, we sought to identify potential differences in OSA with and without the effect of BMI. Thus, we performed gene-based and gene-set MAGMA (v1.08) and eQTL mapping as implemented in FUMA for OSA_mtCOJO_BMI_. Details for these analyses, as well as MAGMA’s Tissue Expression Analysis, LDSC-SEG, and SMR are available in the **Supplementary Note**. As done for the primary OSA meta-analysis, investigated the genes prioritised in the gene discovery phase via spatial transcriptomics analyses using gsMap (v1.71.2)^37^. We filtered our results for those involving genes that were replicated from the primary OSA analysis and had a strict Pearson correlation coefficient (PCC) between the modelled expression and true expression (PCC > 0.70).

We performed Protein–protein interaction (PPI) network analysis using the STRING database v12.0^38^ to evaluate potential biological interactions among genes prioritised by gsMap (PCC > 0.70) in both the European OSA meta-analysis and the BMI-conditioned European OSA analysis. High-confidence interactions (STRING combined score ≥ 0.7) were retained for network construction. Functional modules within the network were identified using the Markov Cluster Algorithm (MCL) with an inflation parameter of 3. Network enrichment was assessed using STRING’s built-in enrichment test, which compares the observed number of interactions with the number expected for a random set of proteins of similar size drawn from the genome.

### SNP-based heritability and genetic correlations

We estimated the heritability for OSA using LDSC(v1.0.1)^39^, with heritability on the liability scale based on a 16% population prevalence^40^. LDSC leverages the expected association between LD variant tags and the expected degree of association with a phenotype. Therefore, variants that are in higher LD with others are expected to capture more of the true polygenic signal, and LDSC leverages this relationship to distinguish polygenic heritability from confounding biases, such as population stratification. Using the munge function from LDSC, we processed the GWAS meta-analysis for OSA and conducted LDSC to estimate the variance explained by the SNPs in the GWAS summary statistics.

We leveraged LDSC to conduct genetic correlation analyses between OSA and several complex human phenotypes. These traits included neuropsychiatric disorders, anthropometric measurements, sleep-related traits, lifestyle factors, cardiometabolic traits, cortical brain measurements, as well as intracranial and subcortical brain volumes. Further details for the GWAS summary statistics for these complex human traits are available in **Supplementary Table 45**. Details on trait selection for this analysis are available in the **Supplementary Note**. We accounted for multiple testing using Bonferroni correction (0.05/228 (total number of genetic correlation tests) = 2.19 x 10^−04^).

As a conditional analysis, we sought to identify potential differences in OSA with and without the effect of BMI. Therefore, we estimated the genetic correlation between OSA and OSA_mtCOJO_BMI_. In addition, we also estimated genetic correlations between OSA, removing the effect of BMI, and complex human traits, ultimately providing genetic correlation results for OSA and OSA_mtCOJO_BMI_ with complex human phenotypes. We accounted for multiple testing using Bonferroni correction (0.05/228 (total number of genetic correlation tests) = 2.19 x 10^−04^). We also estimated genetic correlations among the cohorts of European ancestry that were included in the meta-analysis.

### Colocalisation

Colocalisation using the GWAS-PW (v.0.3.6) method^41^ to identify segments of the genome with genetic variants influencing the etiology of OSA and also another human complex trait is described in detail in the **Supplementary Note**.

### Potential causal genetic effects

We leveraged the Latent Heritable Confounding Mendelian Randomisation (LHC-MR; v0.0.0.9000)^42^ method in R (v4.3.1) to investigate potential causal genetic effects between OSA and the complex human phenotypes with a statistically significant genetic correlation after Bonferroni multiple-testing correction. Details for LHC-MR are available in the **Supplementary Note**. We implemented a Bonferroni multiple testing correction (0.05/34 (total number of LHC-MR tests in the present study) = 1.47 X 10^−03^).

### Polygenic scores estimation and association analyses

We leveraged GWAS summary statistics for OSA to derive PRS for OSA and tested their predictive capability on related outcomes in samples of diverse ancestral backgrounds, including All Of Us^43^ and the Guangzhou Biobank Cohort Study (GBCS)^44^, with full details provided below. Across cohorts, we derived OSA PRS using *SBayesRC (v2.2)*^45^ with the Genome-wide Complex Trait Bayesian (GCTB; v2.0) software tool^46^. PRS were calculated through the multiplication of the multivariate effect size (obtained from *SBayesRC*) times the allelic dosage of the effect allele and summing across all loci for each participant. In addition, to derive the PRS, we only included SNPs passing quality control (MAF > 0.01; call rate > 0.90; and for GBCS an imputation score > 0.60). In the All Of Us cohort, ancestry was defined as having at least 60% composition or greater of a specific ancestry.

Specifically, we leveraged our European ancestry GWAS summary statistics to derive OSA PRS to predict OSA and five features derived from Fitbit data (sleep efficiency^47^, wake after sleep onset (WASO)^48^, number of awakenings^49^, REM sleep percentage^48,50,51^, and fragmentation index^52^) in participants from the All Of Us cohort across ancestries. Fitbit sleep data were obtained from the All of Us Research Program dataset v8, which includes both a bring-your-own-device (BYOD) component and participants enrolled through the WEAR initiative, in which devices were provided directly by the program^53^. To ensure adequate wear duration and data quality, analyses were restricted to participants with at least 21 consecutive days of valid sleep recordings based on timestamped, minute-level sleep stage data. Only nights classified as “main sleep” by the Fitbit algorithm and with full sleep-stage annotation (wake, light, deep, and REM sleep) were retained; nights with only binary “asleep/awake” labels were excluded. Details on sleep stage data handling and harmonisation are available in the **Supplementary Note**.

We note that to evaluate the predictive ability of OSA PRS in European ancestry participants in All Of Us as the target sample, we derived the PRS using a modified version of our original GWAS OSA meta-analysis, in which we left out the All Of Us European ancestry participants from original the meta-analysis. Sample sizes for clinically diagnosed OSA included up to 224,180 European, 77,064 African, 74,036 Admixed American, 9,743 East Asian, 5,362 South Asian, and 1,473 Middle Eastern participants in All Of Us. In contrast, for Fitbit derived measures, sample sizes were substantially reduced, as only a fraction of the participants have these data available. Sample sizes for Fitbit data included up to 10,528 Europeans, 895 Admixed Americans, 713 Africans, 400 East Asians, 135 South Asians, and 32 Middle Eastern participants in All Of Us. Ancestry groups were determined based on individuals who showed 60% or greater ancestry composition for a specific group. A full description of the demographic characteristics and sample sizes for each ancestry is available in **Supplementary Tables 46** and **47**.

We also used OSA PRS derived from the European ancestry GWAS to predict snoring and risk for OSA based on the self-reported 4-item STOP questionnaire^54^ in 2,023 participants of East Asian (Chinese) ancestry from GBCS. The STOP questionnaire was not available in any other cohort leveraged in the present study, so we used it to complement our previous cross-ancestry analyses. In particular, two risk phenotypes for OSA were evaluated for GBCS, (i) having a score of at least two, where one of the points in the score indicates that the patient snores; (ii) having a score of at least three, where one of the points in the score indicates that the patient snores. If a participant had a score of at least two or three, but none of the points represented the presence of snoring, the participant was defined as a control.

Once we derived OSA PRS, we implemented multivariate regressions (logistic for binary outcomes and linear for continuous outcomes) in R for two models. The first one with sex, age, OSA-PRS and the first 10 principal components as covariates to adjust for population stratification. The second one with sex, age, OSA PRS, the first 10 principal components, and BMI as covariates to assess the potential influence of BMI on the predictive ability of the OSA PRS. The BMI covariate used in these models corresponds to a physically examined measure, not a self-reported item. We created figures showing our results in R (v4.3.1) using the *dplyr* package.

## DATA AVAILABILITY

Full GWAS summary statistics for European OSA, European OSA_mtCOJO_BMI_, African-ancestry OSA, and multi-ancestry OSA generated using METAL have been deposited in the NHGRI–EBI GWAS Catalog under accession codes GCST91000712, GCST91000713, GCST91000714 and GCST91000715 (https://www.ebi.ac.uk/gwas/downloads/summary-statistics). Summary statistics for key sensitivity analyses, including European OSA excluding the All of Us cohort (used to derive polygenic scores for All of Us European-ancestry participants), excluding the Australian and Canadian cohorts (OSA EUR no AUS/CAN) and excluding the Million Veteran Program (OSA EUR no MVP), are also available via the GWAS Catalog under accession codes GCST91000716, GCST91000717 and GCST91000718.

Where permitted by data governance agreements, cohort-level summary statistics are shared. For other contributing cohorts, including Mass General Brigham Biobank and the Guangzhou Biobank Cohort Study, as well as the Australian and Canadian cohorts, access to individual-level and cohort-level data requires application through the respective data access processes and data use agreements. Furthermore, summary statistics from contributing individual cohorts such as the Million Veteran Program and FinnGen are publicly available through their respective repositories. Individual-level participant data cannot be made publicly available because of participant consent, privacy requirements and cohort-specific data-use agreements. Work performed using UK Biobank data was conducted under application 25331; UK Biobank data are available to eligible researchers through the UK Biobank access procedure (https://www.ukbiobank.ac.uk/use-our-data/apply-for-access/). Data from the Canadian Longitudinal Study on Aging are available to researchers who meet the criteria for access to de-identified data through the CLSA data access application process (https://www.clsa-elcv.ca/data-access/). This project used data from the National Institutes of Health’s All of Us Research Program’s Controlled Tier Dataset version 8, accessible to authorised researchers on the Researcher Workbench (https://www.researchallofus.org/register/). The GWAS datasets used for genetic-correlation and downstream analyses are identified in Supplementary Table 45. The statistical data underlying the main figures are provided in the Supplementary Tables (Fig. 1, Supplementary Tables 1–3, 6 and 7 and the GWAS Catalog accessions listed above; Fig. 2, Supplementary Tables 22 and 35; Fig. 3, Supplementary Tables 36 and 37; Fig. 4, Supplementary Table 40).

## CODE AVAILABILITY

No custom code was used in this study. Publicly available software tools were used to perform genetic analyses and are referenced throughout the manuscript.

## Notes

### Author Declarations

Our study is based on previously published GWAS summary data and publicly available data for which appropriate site-specific institutional review boards and ethical reviews at local institutions were approved for the use of these data. All participants included in the present study provided written informed consent, and the investigators in the participating studies obtained approval from their institutional review board or equivalent organization. For some cohorts, we conducted the analyses using anonymised individual-level data: AGBD, AGDS, UK Biobank CLSA and AoU. For FinnGen and MVP, we only used publicly available summary data. For the Mass General Brigham Biobank, we also utilised summary data generated as part of a previously published paper (doi: 10.1093/sleep/zsac308).

### Summary of Updates

This version contains revised/updated analyses.

## REFERENCES

1. Slowik, J. M., Sankari, A. & Collen, J. F. Obstructive Sleep Apnea. in StatPearls [Internet] (StatPearls Publishing, 2024).

2. Benjafield, A. V. et al. Estimation of the global prevalence and burden of obstructive sleep apnoea: a literature-based analysis. The Lancet. Respiratory medicine 7, 687 (2019).

3. Malhotra, A. et al. Metrics of sleep apnea severity: beyond the apnea-hypopnea index. Sleep 44, zsab030 (2021).

4. Geer, J. H. & Hilbert, J. Gender Issues in Obstructive Sleep Apnea. The Yale Journal of Biology and Medicine 94, 487 (2021).

5. Romero-Corral, A., Caples, S. M., Lopez-Jimenez, F. & Somers, V. K. Interactions Between Obesity and Obstructive Sleep Apnea: Implications for Treatment. Chest 137, 711 (2010).

6. Campos, A. I. et al. Discovery of genomic loci associated with sleep apnea risk through multi-trait GWAS analysis with snoring. Sleep 46, (2023).

7. Sofer, T. et al. Genome-wide association study of obstructive sleep apnoea in the Million Veteran Program uncovers genetic heterogeneity by sex. EBioMedicine 90, 104536 (2023).

8. Campos, A. I. et al. Insights into the aetiology of snoring from observational and genetic investigations in the UK Biobank. Nature Communications 11, 1–12 (2020).

9. Szily, M. et al. Genetic influences on the onset of obstructive sleep apnoea and daytime sleepiness: a twin study. Respiratory Research 20, 1–6 (2019).

10. Macey, P. M. et al. Brain structural changes in obstructive sleep apnea. Sleep 31, 967–977 (2008).

11. Rosenzweig, I. et al. Changes in Neurocognitive Architecture in Patients with Obstructive Sleep Apnea Treated with Continuous Positive Airway Pressure. EBioMedicine 7, 221–229 (2016).

12. Herrera, C. G. & Tarokh, L. A Thalamocortical Perspective on Sleep Spindle Alterations in Neurodevelopmental Disorders. Current Sleep Medicine Reports 10, 103 (2024).

13. Mölle, M., Bergmann, T. O., Marshall, L. & Born, J. Fast and Slow Spindles during the Sleep Slow Oscillation: Disparate Coalescence and Engagement in Memory Processing. Sleep 34, 1411–1421 (2011).

14. Boxer, E. E. & Aoto, J. Neurexins and their ligands at inhibitory synapses. Frontiers in Synaptic Neuroscience 14, 1087238 (2022).

15. Buraei, Z. & Yang, J. The β subunit of voltage-gated Ca2+ channels. Physiological Reviews 90, 1461–1506 (2010).

16. Bailey, C. S., Moldenhauer, H. J., Park, S. M., Keros, S. & Meredith, A. L. KCNMA1-linked channelopathy. The Journal of General Physiology 151, 1173 (2019).

17. Perea, G. et al. Activity-dependent switch of GABAergic inhibition into glutamatergic excitation in astrocyte-neuron networks. eLife 5, e20362 (2016).

18. Oğuztürk, Ö., Ekici, M., Çimen, D., Ekici, A. & Senturk, E. Attention Deficit/Hyperactivity Disorder in Adults with Sleep Apnea. Journal of Clinical Psychology in Medical Settings 20, 234 (2012).

19. Jehan, S. et al. Depression, Obstructive Sleep Apnea and Psychosocial Health. Sleep medicine and disorders : international journal 1, 00012 (2017).

20. Cubillos-Zapata, C. et al. Differential effect of intermittent hypoxia and sleep fragmentation on PD-1/PD-L1 upregulation. Sleep 43, zsz285 (2019).

21. Sforza, E. & Roche, F. Chronic intermittent hypoxia and obstructive sleep apnea: an experimental and clinical approach. Hypoxia 4, 99 (2016).

22. Ip, M. S. et al. Obstructive sleep apnea is independently associated with insulin resistance. American journal of respiratory and critical care medicine 165, 670–676 (2002).

23. Leung, R. S. Sleep-disordered breathing: autonomic mechanisms and arrhythmias. Progress in cardiovascular diseases 51, 324–338 (2009).

24. Shechter, A. Obstructive sleep apnea and energy balance regulation: A systematic review. Sleep medicine reviews 34, 59–69 (2017).

25. Deng, H. et al. Association of adiposity with risk of obstructive sleep apnea: a population-based study. BMC Public Health 23, 1–12 (2023).

26. Schwartz, A. R. et al. Obesity and Obstructive Sleep Apnea: Pathogenic Mechanisms and Therapeutic Approaches. Proceedings of the American Thoracic Society 5, 185 (2008).

27. Cowan, D. C. & Livingston, E. Obstructive Sleep Apnoea Syndrome and Weight Loss: Review. Sleep Disorders 2012, 163296 (2012).

28. Zhu, Z. et al. Causal associations between risk factors and common diseases inferred from GWAS summary data. Nature Communications 9, 1–12 (2018).

## METHODS-ONLY REFERENCES

29. Zhou, W. et al. Efficiently controlling for case-control imbalance and sample relatedness in large-scale genetic association studies. Nature Genetics 50, 1335–1341 (2018).

30. Mbatchou, J. et al. Computationally efficient whole-genome regression for quantitative and binary traits. Nature Genetics 53, 1097–1103 (2021).

31. Willer, C. J., Li, Y. & Abecasis, G. R. METAL: fast and efficient meta-analysis of genomewide association scans. Bioinformatics 26, 2190 (2010).

32. Purcell, S. et al. PLINK: A Tool Set for Whole-Genome Association and Population-Based Linkage Analyses. American Journal of Human Genetics 81, 559 (2007).

33. Yang, J. et al. Conditional and joint multiple-SNP analysis of GWAS summary statistics identifies additional variants influencing complex traits. Nature genetics 44, 369–375 (2012).

34. Loos, R. J. F. & Yeo, G. S. H. The bigger picture of FTO—the first GWAS-identified obesity gene. Nature Reviews Endocrinology 10, 51–61 (2013).

35. Huang, J. et al. Genomics and phenomics of body mass index reveals a complex disease network. Nature Communications 13, 1–10 (2022).

36. Brown, B. C., Asian Genetic Epidemiology Network Type 2 Diabetes Consortium, Ye, C. J., Price, A. L. & Zaitlen, N. Transethnic Genetic-Correlation Estimates from Summary Statistics. American Journal of Human Genetics 99, 76 (2016).

37. Song, L., Chen, W., Hou, J., Guo, M. & Yang, J. Spatially resolved mapping of cells associated with human complex traits. Nature 641, 932–941 (2025).

38. Szklarczyk, D. et al. The STRING database in 2023: protein-protein association networks and functional enrichment analyses for any sequenced genome of interest. Nucleic Acids Res. 51, D638–D646 (2023).

39. Bulik-Sullivan, B. K. et al. LD Score regression distinguishes confounding from polygenicity in genome-wide association studies. Nature genetics 47, 291–295 (2015).

40. Zasadzińska-Stempniak, K., Zajączkiewicz, H. & Kukwa, A. Prevalence of Obstructive Sleep Apnea in the Young Adult Population: A Systematic Review. Journal of clinical medicine 13, (2024).

41. Pickrell, J. K. et al. Detection and interpretation of shared genetic influences on 42 human traits. Nature genetics 48, 709–717 (2016).

42. Darrous, L., Mounier, N. & Kutalik, Z. Simultaneous estimation of bi-directional causal effects and heritable confounding from GWAS summary statistics. Nature communications 12, 7274 (2021).

43. All of Us Research Program Genomics Investigators. Genomic data in the All of Us Research Program. Nature 627, 340–346 (2024).

44. Jiang, C. Q. et al. An overview of the Guangzhou biobank cohort study-cardiovascular disease subcohort (GBCS-CVD): a platform for multidisciplinary collaboration. Journal of human hypertension 24, 139–150 (2010).

45. Zheng, Z. et al. Leveraging functional genomic annotations and genome coverage to improve polygenic prediction of complex traits within and between ancestries. Nature genetics 56, 767–777 (2024).

46. Zeng, J. et al. Signatures of negative selection in the genetic architecture of human complex traits. Nature Genetics 50, 746–753 (2018).

47. Adekolu, O. & Zinchuk, A. Sleep Deficiency in Obstructive Sleep Apnea. Clinics in chest medicine 43, 353 (2022).

48. Park, M., Senel, G. B., Modi, H., Jain, V. & DelRosso, L. M. Combined impact of obstructive sleep apnea and periodic limb movements on sleep parameters. Sleep medicine 129, (2025).

49. Krakow, B., Romero, E., Ulibarri, V. A. & Kikta, S. Prospective Assessment of Nocturnal Awakenings in a Case Series of Treatment-Seeking Chronic Insomnia Patients: A Pilot Study of Subjective and Objective Causes. Sleep 35, 1685 (2012).

50. Bonsignore, M. R. et al. REM sleep obstructive sleep apnoea. European Respiratory Review 33, 230166 (2024).

51. Rishi, A. R. & Rishi, M. A. Rapid eye movement related obstructive sleep apnea: Where do we stand? Respiratory Investigation 59, 589–595 (2021).

52. Staats, R. et al. The Importance of Sleep Fragmentation on the Hemodynamic Dipping in Obstructive Sleep Apnea Patients. Frontiers in Physiology 11, 104 (2020).

53. Bailey, C. P. et al. Fitbit Physical Activity and Sleep Data in the All of Us Research Program: Data Exploration and Processing Considerations for Research. Medicine and science in sports and exercise 10.1249/MSS.0000000000003804 (2025) doi:10.1249/MSS.0000000000003804.

54. Patel, D. et al. Validation of the STOP questionnaire as a screening tool for OSA among different populations: a systematic review and meta-regression analysis. Journal of Clinical Sleep Medicine : JCSM : Official Publication of the American Academy of Sleep Medicine 18, 1441 (2022).

