## Supplementary Material for "Genome-wide analyses identify 147 loci associated with obstructive sleep apnea"

### Supplementary results

***GWAS Meta-Analysis Sensitivity Analyses in Europeans***

We performed three sensitivity analyses to evaluate the robustness of the primary meta-analysis to phenotype definition, cohort composition, and potential misclassification of undiagnosed OSA. First, to assess the impact of phenotype heterogeneity arising from symptom-based self-report definitions of OSA, we excluded the Australian and Canadian cohorts. Results were highly concordant with the primary meta-analysis (**Supplementary Table 1**). Specifically, 146 of the 147 genome-wide significant loci identified in the primary analysis remained genome-wide significant after cohort exclusion, with all lead variants retaining the same direction of effect and effect size estimates remaining nearly identical across loci. The single locus that fell marginally below genome-wide significance (rs2879031) showed minimal change in effect size, consistent with the modest reduction in total sample size.

Second, to ensure results were not driven by any single large contributing cohort, we conducted a leave-one-out meta-analysis excluding the Million Veteran Program (MVP). Effect size estimates were highly concordant with those from the full meta-analysis (**Supplementary Table 1**), with preservation of effect direction across all lead variants and the vast majority of genome-wide significant loci remaining statistically significant.

Third, to evaluate the robustness of our findings to potential misclassification of undiagnosed OSA among controls in the UK Biobank, we performed a sensitivity analysis using a more stringent definition for the control sample, representing a ~2% reduction in the control sample (*see Methods*). Results were highly concordant with those from our original UK Biobank GWAS, with consistent magnitude and direction of effect sizes across loci (**Supplementary Table 1**). Manhattan and quantile–quantile plots for all sensitivity analyses are available in **Supplementary Figures 18 - 23**.

***GWAS in African Ancestry***

We examined whether African-ancestry lead variants were also associated in the European meta-analysis. Two (rs1558902 and rs9825254) of the six African-ancestry loci were genome-wide significant in Europeans with concordant effect directions, while other loci showed weaker (rs1164548 and rs146615151) or no evidence of association (rs62502796 and rs111744602) and were not in linkage disequilibrium with European lead variants. The variant rs111744602 lies within an intron of the long non-coding RNA gene *ZFHX3-AS1*, whereas rs62502796 is located within the intronic regions of two long non-coding RNA transcripts on chromosome eight.

***Functional Annotation and Gene Prioritisation***

**Gene discovery and pathway analysis.** Gene-based association tests using MAGMA and fastBAT yielded consistent findings (MAGMA $P < 2.65 \times 10^{-06}$; fastBAT $P < 2.04 \times 10^{-06}$; *see Methods*), implicating hundreds of genes after multiple testing correction (e.g., *CADM2, FTO, ETV5, AUTS2, SCAPER, APOE, ATXN2L*; *see* **Supplementary Tables 9 and 10** for full lists). Subsequent gene-set analysis revealed enrichment in 17 pathways, including neurogenesis, axon development, and neuron development (**Supplementary Tables 11 and 12**).

**Tissue and cell-type specificity.** We next performed three complementary analyses, MAGMA tissue-expression analysis, LDSC-SEG, and eQTL mapping, to investigate tissue specificity. These analyses converged to show significant enrichment of OSA-associated gene expression in the brain, particularly in the cerebellum, frontal cortex, and subcortical basal ganglia structures (e.g., nucleus accumbens, caudate nucleus) ($P < 9.26 \times 10^{-04}$; *see Methods*; **Supplementary Tables 13-15**). Integrating the OSA GWAS with GTEx v8 brain eQTL data further supported these findings, identifying numerous genes whose expression in these brain regions is associated with OSA. Across all 13 brain tissues tested, six genes were ubiquitously implicated: *LRRC37A2, AS3MT, TTC12, LRRC37A, GTF2IRD2,* and *ZSCAN31* (**Supplementary Tables 16 and 17**).

**Conditional analysis removing BMI effects.** Tissue-expression analyses showed a marked attenuation of tissue-specific enrichment signals. In MAGMA general tissue and specific tissue analyses, no tissues remained significantly enriched after correction for multiple testing (**Supplementary Tables 27** and **28)**. Nonetheless, we observed nominally significant associations ($P < 0.05$), including enrichment in structural and peripheral tissues such as uterus, skeletal muscle, cervix uteri, and ovary in MAGMA's general tissue analysis, as well as cerebellum, cerebellar hemisphere, tibial artery, uterus, and skeletal muscle in MAGMA's specific tissue analysis. Integrating the $OSA_{mtCOJO\_BMI}$ GWAS with GTEx v8 brain eQTL data uncovered numerous genes expressed in the cortex and across subcortical brain structures (**Supplementary Tables 29** and **30)**. LDSC-SEG analyses conducted on the same brain and peripheral tissues that were significantly enriched in the primary OSA meta-analysis yielded no statistically significant enrichments following BMI conditioning (**Supplementary Table 31**). Summary-based Mendelian Randomisation (SMR), consistent with our OSA primary results, identified *SCAPER* as a potential causal gene in the cerebellum ($P < 7.64\times10^{-07}$; **Supplementary Table 32**), along with numerous other genes of nominal statistical significance ($P < 0.05$) across cortical and subcortical prioritised tissues (**Supplementary Table 33**).

#### *Genetic Overlap and Causality with Other Complex Traits*

**Colocalisation.** Further analysis with GWAS-PW identified 544 unique genomic segments jointly influencing OSA and at least one other phenotype (**Supplementary Table 42**). Cardiometabolic traits, such as BMI ($n$ = 344), type 2 diabetes ($n$ = 198), and HDL cholesterol ($n$ = 100), shared the largest number of genomic segments ($n$) with OSA, with 49 genomic segments being common for the aforementioned traits. Similarly, neuropsychiatric traits, including major depressive disorder ($n$ = 87), multisite chronic pain ($n$ = 50), neuroticism score ($n$ = 42), ADHD ($n$ = 39), and schizophrenia ($n$ = 34), shared a considerable number of genomic segments with OSA.

### Supplementary discussion

Our gene-level analyses indicate that a substantial proportion of the genetic signal for OSA is mediated through BMI-related pathways. Following BMI conditioning, the number of associated genes was markedly reduced, and several genes highlighted in the primary analysis, such as *CADM2*, *ETV5*, and *AUTS2*, were no longer statistically significant after multiple testing correction, consistent with mediation by adiposity and/or shared pleiotropic effects. The attenuation of *FTO*, while not complete, further supports a contribution of obesity-related biology to OSA risk. Notably, *MSRB3* remained associated after BMI conditioning, suggesting a potential role for oxidative stress or mitochondrial function independent of BMI, which are processes implicated in neuromuscular and inflammatory components of sleep-disordered breathing[1–3]. Together, these results suggest that while much of the gene-level signal reflects adiposity-driven mechanisms, a smaller subset of loci may contribute to OSA risk through adiposity-independent pathways.

Tissue-specific analyses of the primary OSA GWAS suggested enrichment across multiple brain regions, consistent with neural regulation of breathing and arousal[4]. However, these tissue-level signals were attenuated following BMI conditioning, in which no tissues remained significantly enriched after multiple testing correction in MAGMA general or specific tissue analyses, and LDSC-SEG analyses likewise showed no statistically significant enrichment. Nominal associations ($P < 0.05$) were observed in a mixture of structural and peripheral tissues, including skeletal muscle, uterus, and cerebellar regions. This pattern suggests that the broad, tissue-level brain enrichment observed in the primary analysis is largely driven by BMI-related genetic effects. These observations are consistent with the known role of hypothalamic and basal ganglia circuits in energy balance and adiposity[5], suggesting that the apparent brain-wide enrichment in the primary unadjusted analysis is at least partly attributable to BMI-related genetic effects rather than OSA-specific neurobiology. Nonetheless, fine-grained spatial analyses indicate that a subset of potential BMI-independent associations persists within specific cortical circuits (*see discussion on spatial mapping below*), highlighting the importance of higher-resolution approaches when evaluating potential residual neurobiological contributions to OSA risk.

Phenotypic heterogeneity across cohorts is an important consideration in large-scale meta-analyses of complex traits. In this study, OSA was defined using both clinically ascertained diagnoses (ICD-based or electronic health record–derived phenotypes) and structured self-reported questionnaire data. To evaluate the impact of this heterogeneity, we estimated genetic correlations among all OSA cohorts included in the meta-analysis. These analyses revealed predominantly positive and moderate-to-high genetic correlations, indicating substantial shared genetic architecture across cohorts despite differences in ascertainment strategies. Although some correlations, particularly those involving smaller self-report–based cohorts, did not reach nominal statistical significance, this was largely attributable to reduced statistical power and large standard errors rather than discordant genetic effects. Consistent with this interpretation, point estimates were overall positive and comparable in magnitude across cohorts. A small number of genetic correlation estimates exceeded 1, a known artifact of LD Score regression that reflects sampling variance in underpowered samples rather than differences in genetic architecture[6].

Supplementary sensitivity analyses further supported the robustness of the $OSA_{mtCOJO\_BMI}$ findings. Across the 147 independent genome-wide loci identified for the primary OSA meta-analysis, our $OSA_{mtCOJO\_BMI}$ analyses resulted in a consistent attenuation of effect sizes rather than widespread loss or change in the direction of effect sizes, with those loci showing stronger attenuation exhibiting stronger associations with BMI. Directionality analyses across the 39 $OSA_{mtCOJO\_BMI}$ independent loci revealed a high degree of concordance between unconditioned and BMI-conditioned OSA effects, with directionality reversals occurring at only a few loci. Genetic variants exhibiting collider patterns (defined by opposing effect size directions between BMI and $OSA_{mtCOJO\_BMI}$ at genome-wide BMI statistical significance) were rare and largely confined to loci with extremely strong BMI associations. These findings suggest that while a minority of loci may remain sensitive to residual pleiotropy or conditioning artefacts, the majority of $OSA_{mtCOJO\_BMI}$ associations reflect stable genetic effects not driven solely by adiposity. Together, these results reinforce the interpretation that OSA risk arises from both BMI-mediated and BMI-independent biological mechanisms.

Additional sensitivity analyses further support the robustness of our findings. Exclusion of cohorts with symptom-based self-report definitions of OSA (the Australian cohorts and CLSA) had minimal impact on the overall genetic architecture, with nearly all genome-wide significant loci retained, consistent effect directions across lead variants, and negligible changes in effect size estimates. Similarly, a leave-one-out meta-analysis excluding the largest contributing cohort (MVP) yielded highly concordant results, indicating that the observed associations are not driven by any single cohort.

We acknowledge this study has several limitations. First, OSA is frequently underdiagnosed[7,8], creating a risk of misclassifying cases within control groups. Second, reliance on self-report questionnaires in some cohorts, as opposed to clinical ICD-10 or SNOMED codes, could lead to the inadvertent inclusion of a small number of central sleep apnoea cases. To mitigate both issues, we applied stringent, multi-item criteria for case-control definitions in all self-report cohorts (see *Methods*), which minimises these potential sources of misclassification. Furthermore, given that the population prevalence of central sleep apnoea is low (<1%) and typically skews older than our study's predominant age group[9], any minor inclusion of central sleep apnoea is unlikely to have substantially influenced our findings.

In the discovery phase, our GWAS meta-analysis only included participants of European ancestry. Thus, the genome-wide loci reported here are only representative for individuals of European ancestry until confirmed in samples of other ancestral backgrounds, with the exception of those genetic loci identified in the independent meta-analysis for individuals of African ancestry, which doubled those reported in a previous African GWAS[10]. We note that the multi-ancestry meta-analysis yielded fewer independent loci than the European-only analysis, which likely reflects differences in linkage disequilibrium structure across ancestries and the use of a multi-ancestry 1000 Genomes reference panel (~2,500 individuals) for locus definition, resulting in more conservative consolidation of association signals. Regarding the estimation of potential causal genetic effects, we detected slight evidence of horizontal pleiotropy for some cardiometabolic phenotypes. However, LHC-MR models both sample overlap and correlated pleiotropy, providing putative causal estimates considering these factors. While not all genetic overlap between OSA and another complex human phenotype may be causal, the strength and robustness of the remaining causal components suggest it is unlikely that our findings are explained by pleiotropy alone. Similarly, we note that while European-derived OSA PRS showed encouraging predictive power across ancestries, the clinical utility of these scores may ultimately be greatest when derived from ancestry-specific GWAS data. These findings highlight the relevance and potential of genomics for clinical screening of patients at high risk for OSA. Lastly, replication of our findings in GWAS of polysomnography-derived traits (e.g., apnea–hypopnea index or oxygen desaturation) is currently limited by the lack of publicly available, well-powered datasets, representing an important direction for future research as such resources become available.

### Supplementary figures

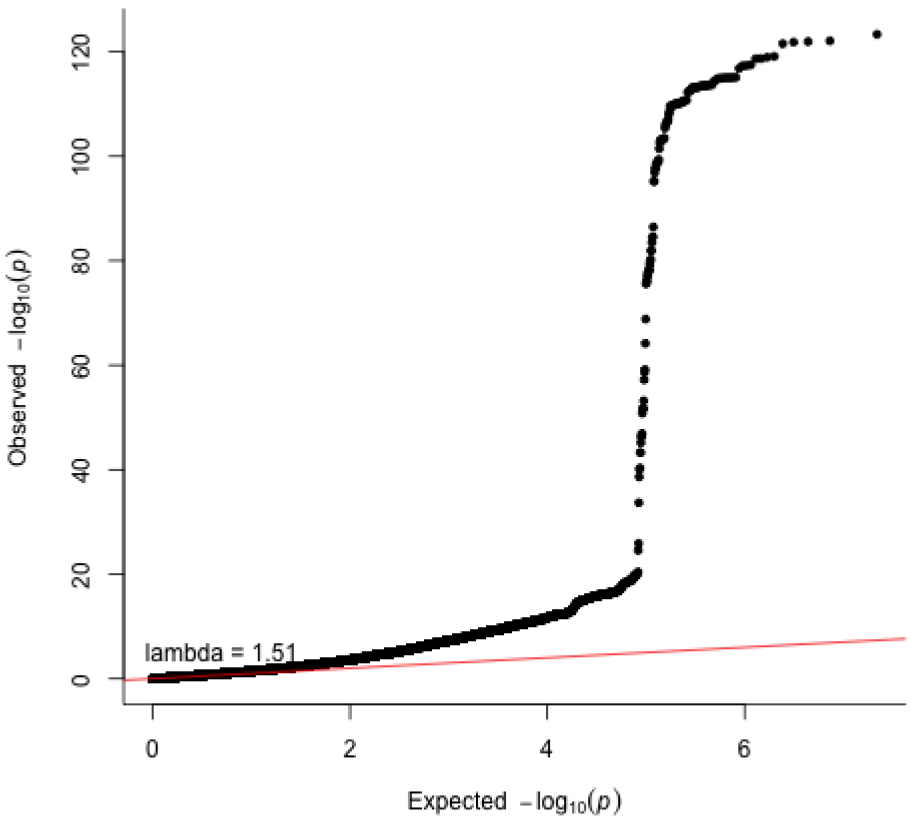


**Supplementary Figure 1. Obstructive sleep apnoea European meta-analysis QQ plot (n = 1,491,682; 232,292 cases and 1,259,390 controls).** The p-values referenced here correspond to a two-tailed Z-test as implemented in METAL. Each dot compares the observed and expected −log10(P) values. The red diagonal line indicates the expected distribution under the null hypothesis. λGC denotes the genomic inflation factor.

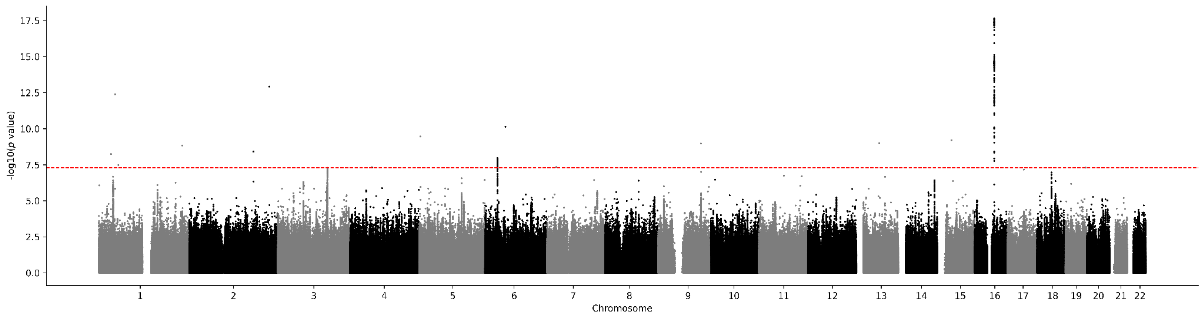


**Supplementary Figure 2. Obstructive sleep apnoea Manhattan plot in AllOfUs.** Results for obstructive sleep apnoea GWAS (n = 225,598; 32,618 cases and 192,980 controls). Genome-wide significance is shown for the common threshold of *p*-value < $5x10^{-8}$ (red dashed line). The p-values referenced here correspond to a two-sided Wald test ($\chi^2_1$) as implemented in REGENIE.

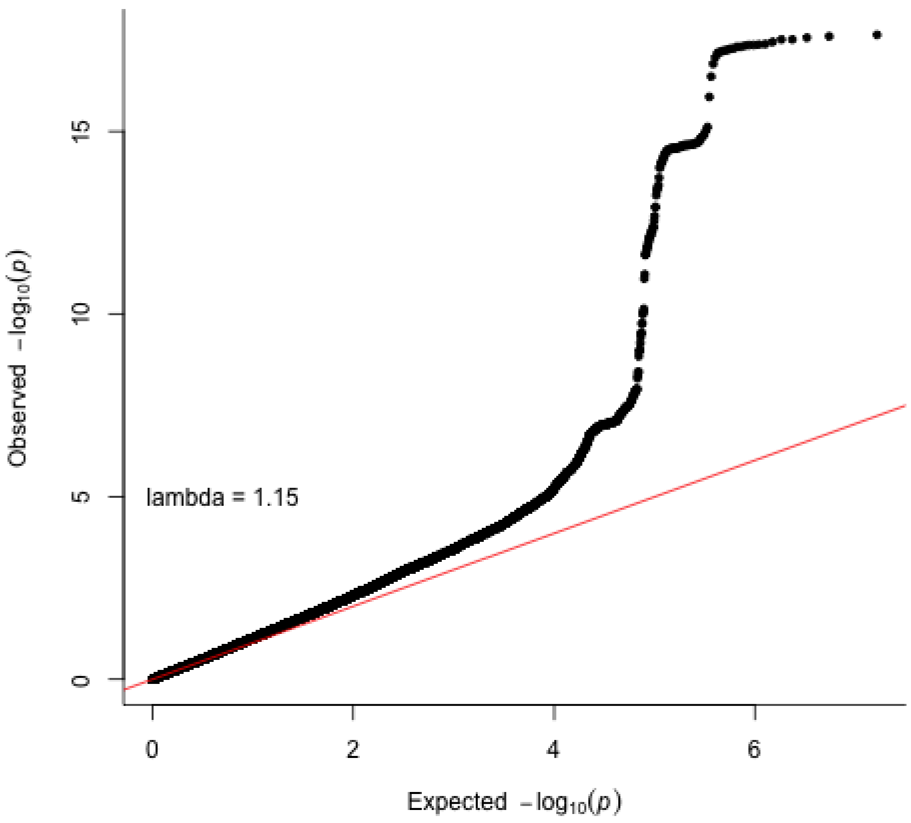


**Supplementary Figure 3. Obstructive sleep apnoea QQ plot in AllOfUs (n = 225,598; 32,618 cases and 192,980 controls).** The p-values referenced here correspond to a two-sided Wald test ($\chi^2_1$) as implemented in REGENIE. Each dot compares the observed and expected −log10(P) values. The red diagonal line indicates the expected distribution under the null hypothesis. λGC denotes the genomic inflation factor.

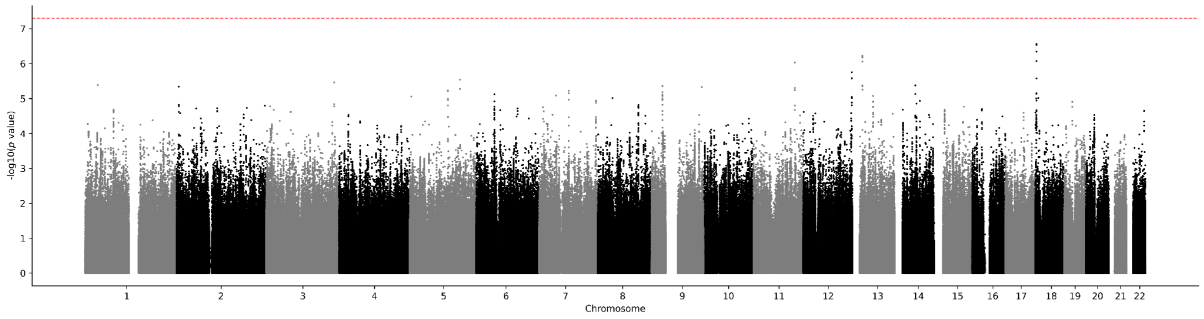


**Supplementary Figure 4. Obstructive sleep apnoea Manhattan plot in Australian cohorts.** Results for obstructive sleep apnoea GWAS (n = 9,636; 1,247 cases and 8,389 controls). Genome-wide significance is shown for the common threshold of *p*-value < $5x10^{-8}$ (red dashed line). The p-values referenced here correspond to a two-sided **score test** ($\chi^2{}_1$) as implemented in SAIGE's mixed-model framework.

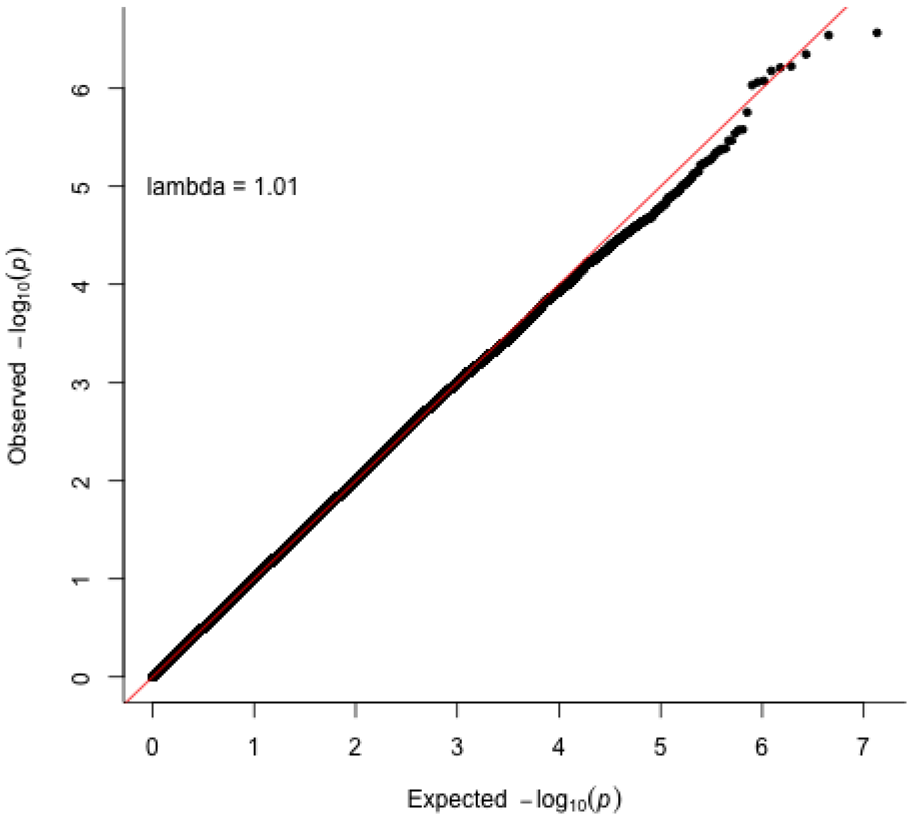


**Supplementary Figure 5. Obstructive sleep apnoea QQ plot in Australian cohorts (n = 9,636; 1,247 cases and 8,389 controls).** The p-values referenced here correspond to a two-sided score test ($\chi^2{}_1$) as implemented in SAIGE's mixed-model framework. Each dot compares the observed and expected −log10(P) values. The red diagonal line indicates the expected distribution under the null hypothesis. λGC denotes the genomic inflation factor.

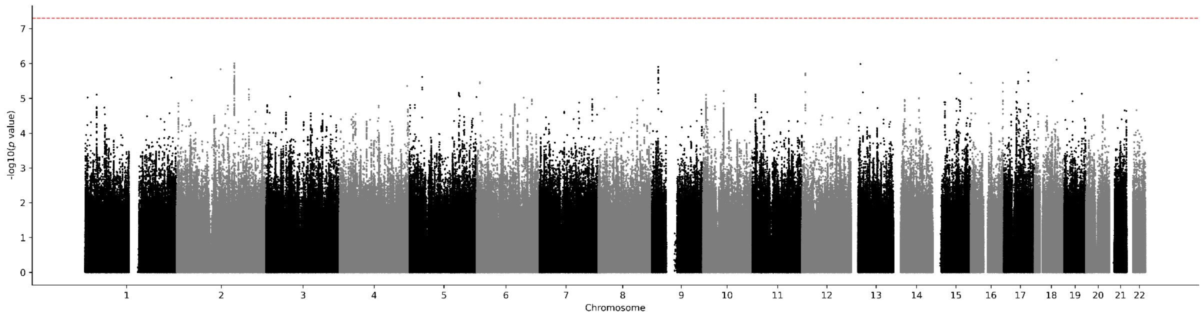


**Supplementary Figure 6. Obstructive sleep apnoea Manhattan plot in CLSA.** Results for obstructive sleep apnoea GWAS (n = 15,587; 4,151 cases and 11,436 controls). Genome-wide significance is shown for the common threshold of *p*-value < $5x10^{-8}$ (red dashed line). The p-values referenced here correspond to a two-sided Wald test ($\chi^2{}_1$) as implemented in REGENIE.

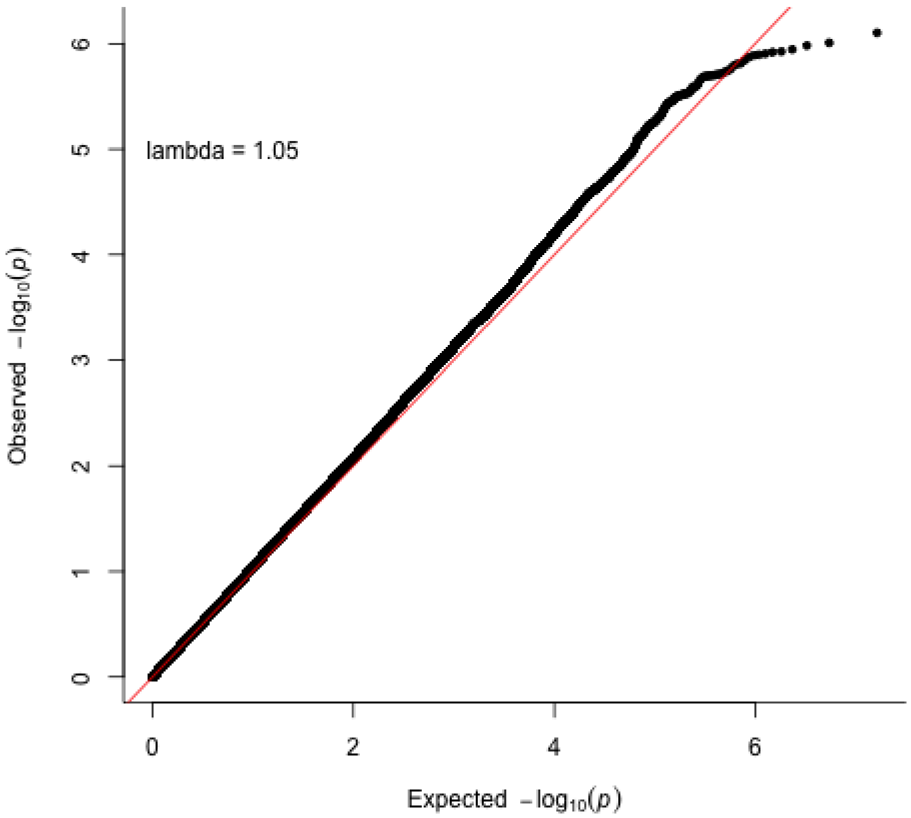


**Supplementary Figure 7. Obstructive sleep apnoea QQ plot in CLSA (n = 15,587; 4,151 cases and 11,436 controls).** The p-values referenced here correspond to a two-sided Wald test ($\chi^2{}_1$) as implemented in REGENIE. Each dot compares the observed and expected –log10(P) values. The red diagonal line indicates the expected distribution under the null hypothesis. λGC denotes the genomic inflation factor.

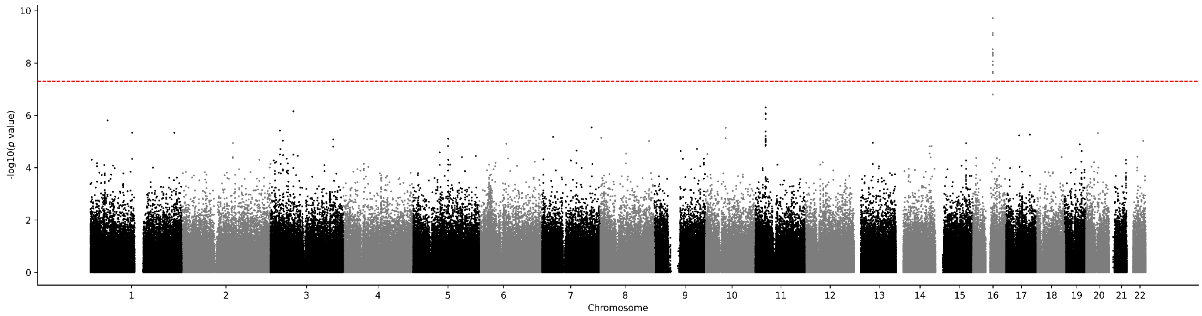


**Supplementary Figure 8. Obstructive sleep apnoea Manhattan plot in UK Biobank.** Results for obstructive sleep apnoea GWAS (n = 270,279; 5,959 cases and 264,320 controls). Genome-wide significance is shown for the common threshold of *p*-value < 5x10$^{-8}$ (red dashed line). The p-values referenced here correspond to a two-sided score test ($\chi^2_1$) as implemented in SAIGE's mixed-model framework.

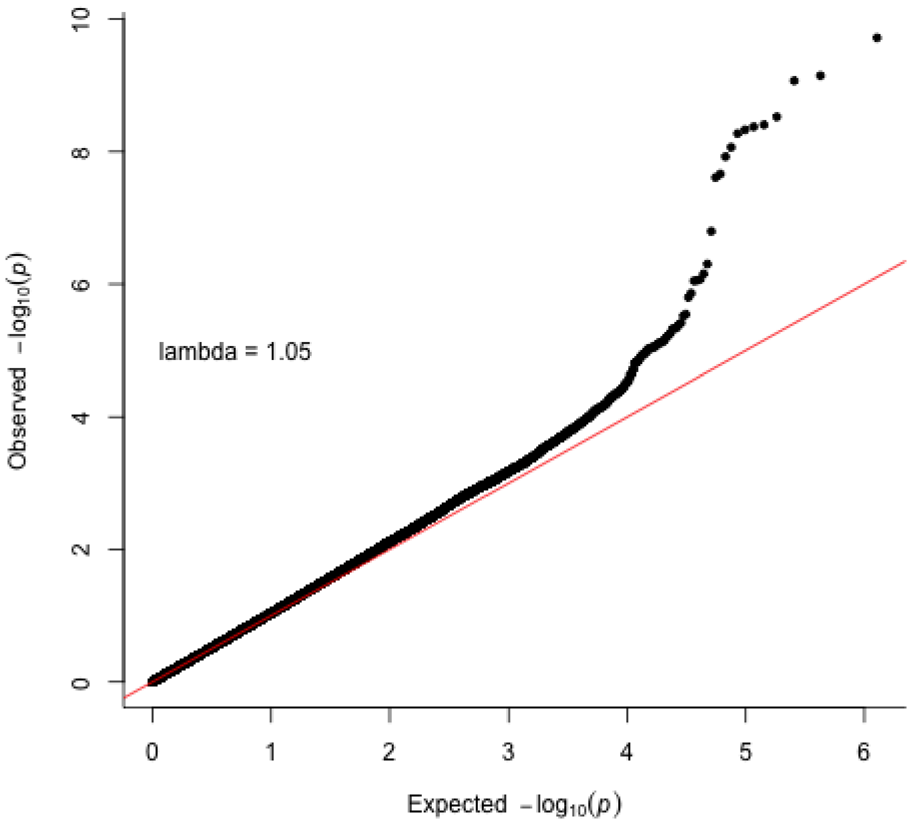


**Supplementary Figure 9. Obstructive sleep apnoea QQ plot in UK Biobank (n = 270,279; 5,959 cases and 264,320 controls).** The p-values referenced here correspond to a two-sided score test ($\chi^2_1$) as implemented in SAIGE's mixed-model framework. Each dot compares the observed and expected −log10(P) values. The red diagonal line indicates the expected distribution under the null hypothesis. λGC denotes the genomic inflation factor.

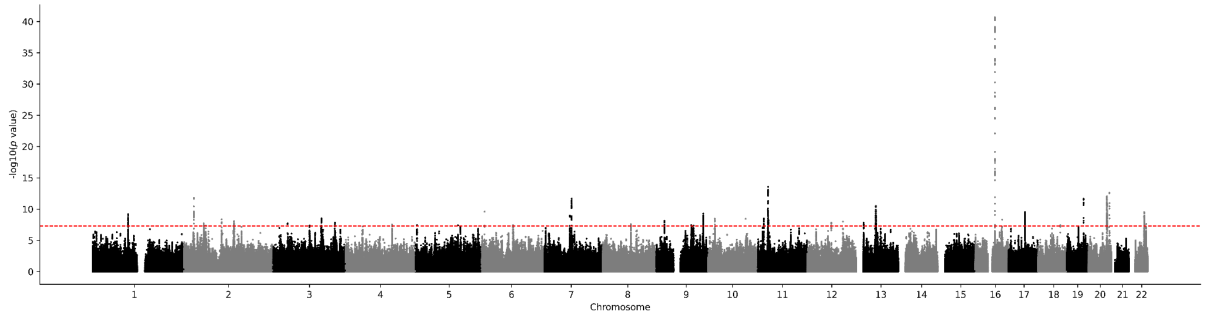


**Supplementary Figure 10. Obstructive sleep apnoea Manhattan plot in FinnGen.** Results for obstructive sleep apnoea GWAS (n = 520,210; 58,520 cases and 461,690 controls). Genome-wide significance is shown for the common threshold of *p*-value < $5x10^{-8}$ (red dashed line). The p-values referenced here correspond to a two-sided score test ($\chi^2{}_1$) as implemented in SAIGE's mixed-model framework.

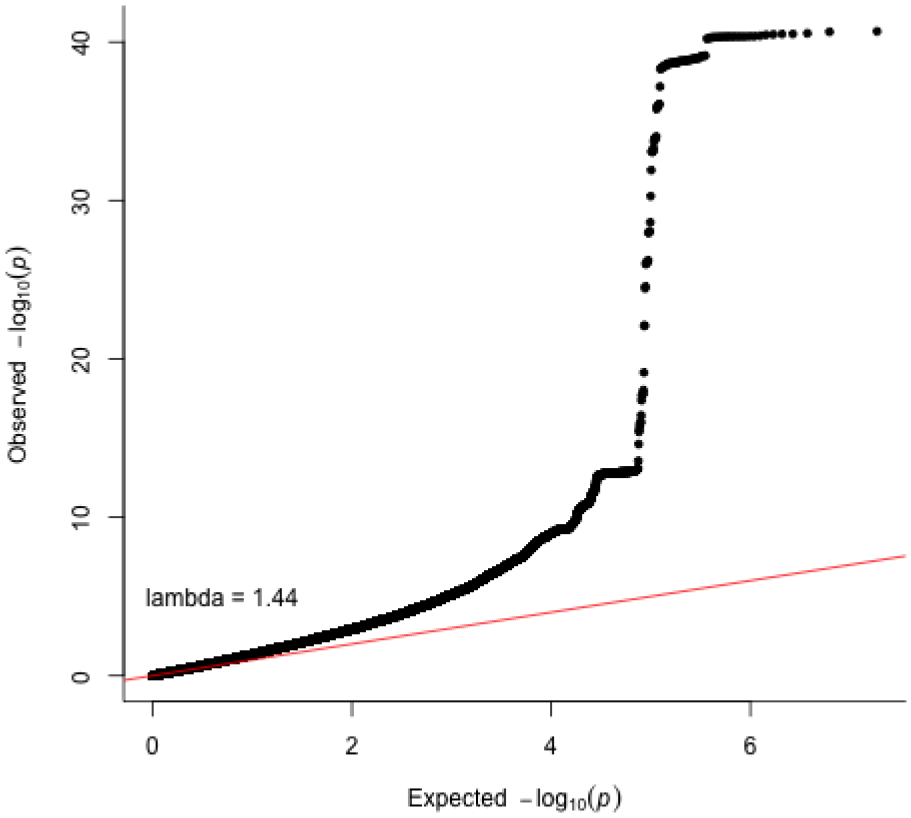


**Supplementary Figure 11. Obstructive sleep apnoea QQ plot in FinnGen (n = 520,210; 58,520 cases and 461,690 controls).** The p-values referenced here correspond to a two-sided score test ($\chi^2{}_1$) as implemented in SAIGE's mixed-model framework. Each dot compares the observed and expected −log10(P) values. The red diagonal line indicates the expected distribution under the null hypothesis. λGC denotes the genomic inflation factor.

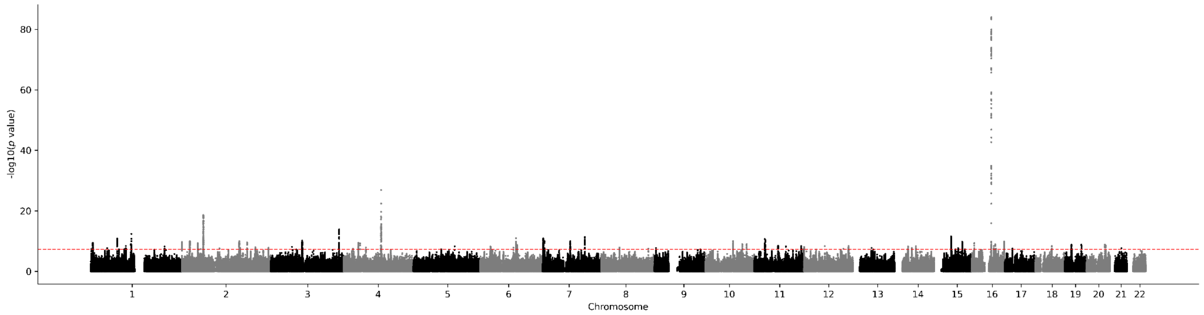


**Supplementary Figure 12. Obstructive sleep apnoea Manhattan plot in MVP.** Results for obstructive sleep apnoea GWAS (n = 430,325; 126,695 cases and 303,630 controls). Genome-wide significance is shown for the common threshold of *p*-value < $5x10^{-8}$ (red dashed line). The p-values referenced here correspond to a two-sided score test ($\chi^2{}_1$) as implemented in SAIGE's mixed-model framework.

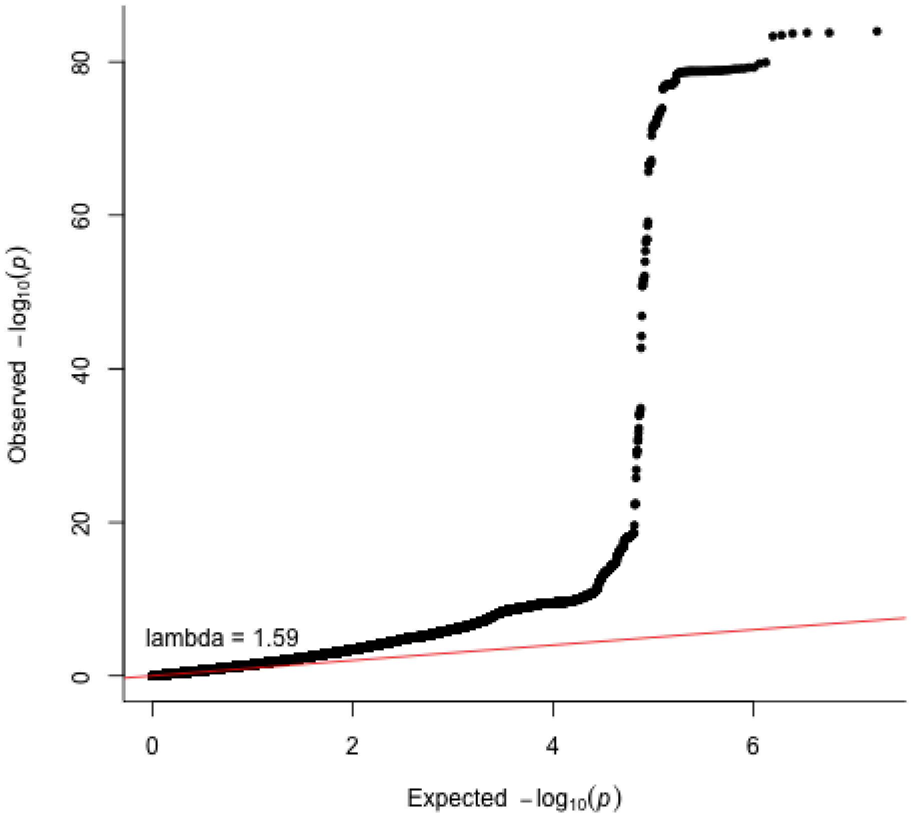


**Supplementary Figure 13. Obstructive sleep apnoea QQ plot in MVP (n = 430,325; 126,695 cases and 303,630 controls).** The p-values referenced here correspond to a two-sided score test ($\chi^2{}_1$) as implemented in SAIGE's mixed-model framework. Each dot compares the observed and expected –log10(P) values. The red diagonal line indicates the expected distribution under the null hypothesis. λGC denotes the genomic inflation factor.

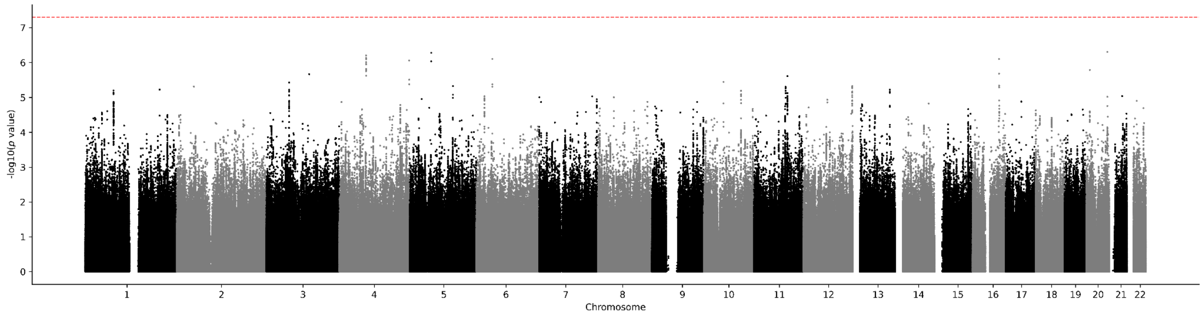


**Supplementary Figure 14. Obstructive sleep apnoea Manhattan plot in Mass General Brigham Biobank.** Results for obstructive sleep apnoea GWAS (n = 20,047; 3,102 cases and 16,945 controls). Genome-wide significance is shown for the common threshold of *p*-value < $5x10^{-8}$ (red dashed line). The p-values referenced here correspond to a two-sided Wald test ($\chi^2_1$) as implemented in PLINK.

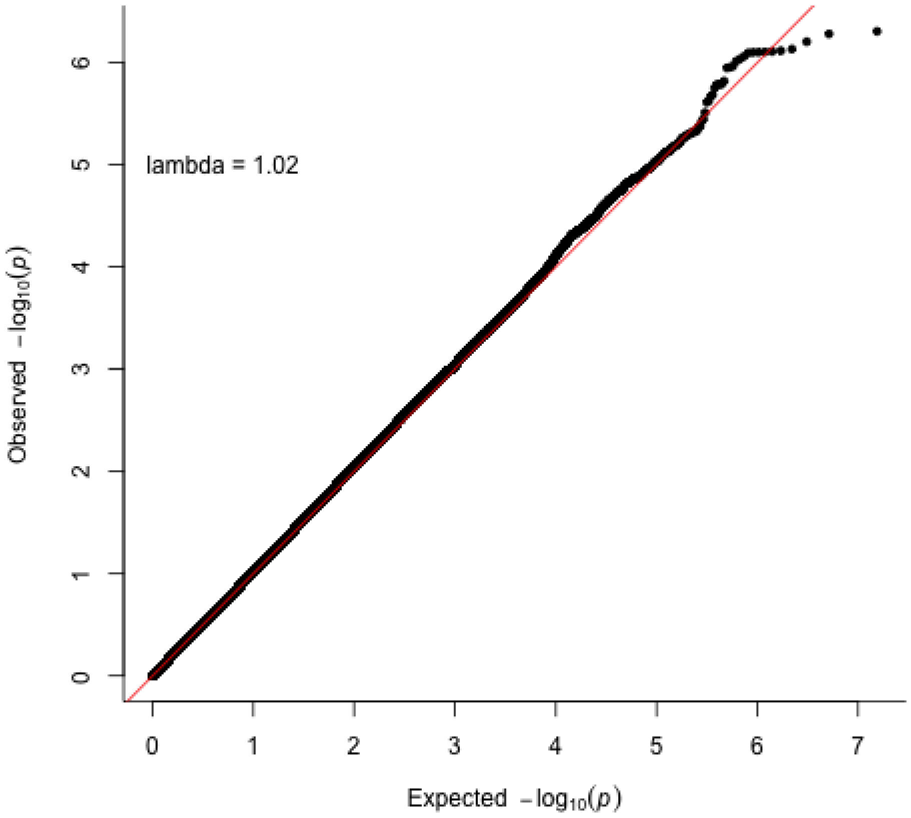


**Supplementary Figure 15. Obstructive sleep apnoea QQ plot in Mass General Brigham Biobank (n = 20,047; 3,102 cases and 16,945 controls).** The p-values referenced here correspond to a two-sided Wald test ($\chi^2_1$) as implemented in PLINK. Each dot compares the observed and expected −log10(P) values. The red diagonal line indicates the expected distribution under the null hypothesis. λGC denotes the genomic inflation factor.

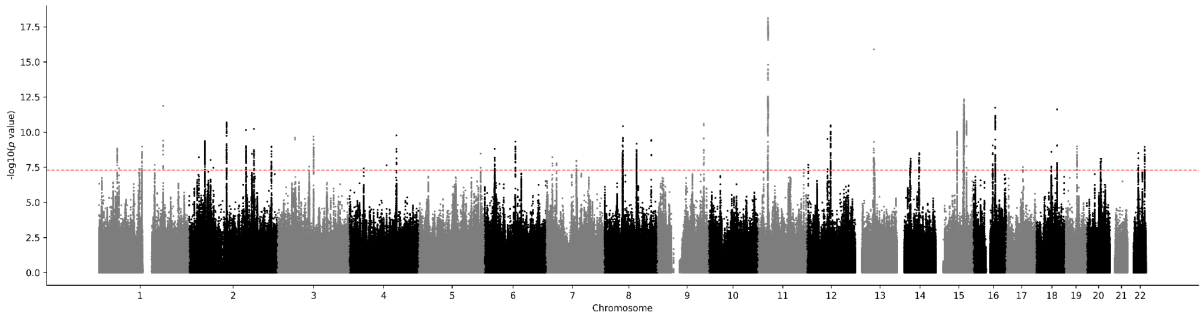


**Supplementary Figure 16. Obstructive sleep apnoea (mtCOJO BMI) Manhattan plot.** Results for obstructive sleep apnoea GWAS (n = 1,491,682; 232,292 cases and 1,259,390 controls). Genome-wide significance is shown for the common threshold of *p*-value < $5x10^{-8}$ (red dashed line). The p-values referenced here correspond to a two-sided Z-test as implemented in mtCOJO.

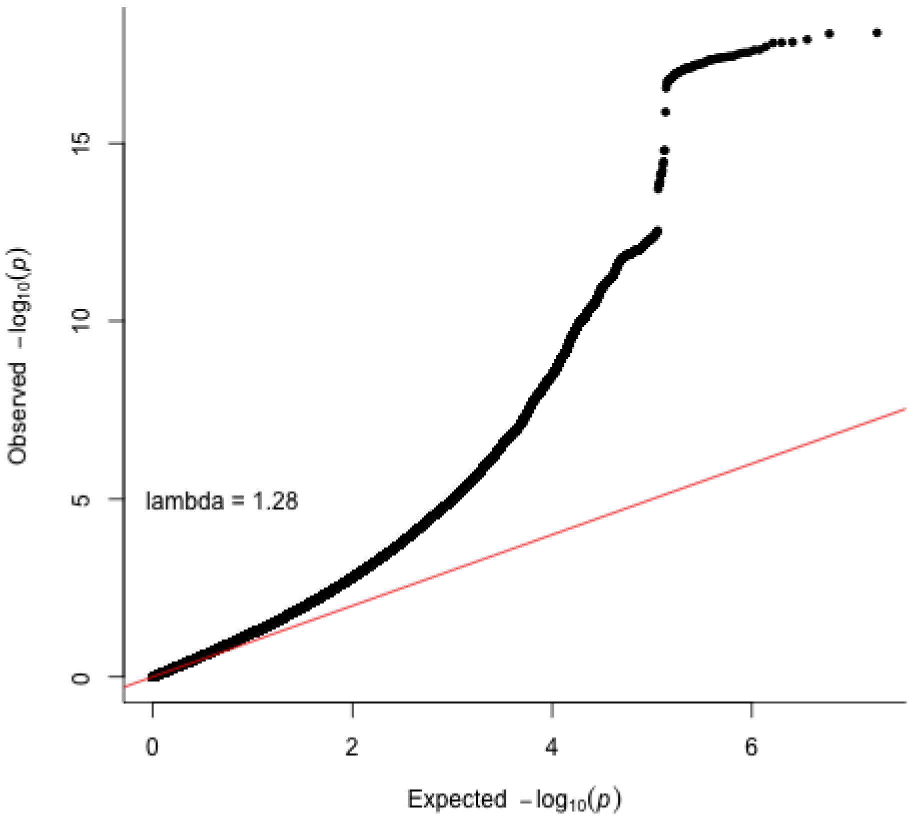


**Supplementary Figure 17. Obstructive sleep apnoea (mtCOJO BMI) meta-analysis QQ plot (n = 1,491,682; 232,292 cases and 1,259,390 controls).** The p-values referenced here correspond to a two-sided Z-test as implemented in mtCOJO. Each dot compares the observed and expected −log10(P) values. The red diagonal line indicates the expected distribution under the null hypothesis. λGC denotes the genomic inflation factor.

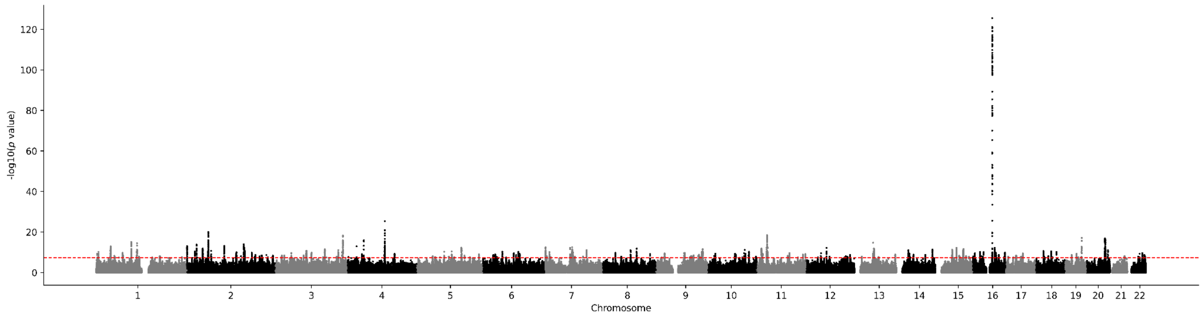


**Supplementary Figure 18. Obstructive sleep apnoea European meta-analysis excluding the Australian cohorts and CLSA Manhattan plot.** Results for obstructive sleep apnoea GWAS (n = 1,466,459; 226,894 cases and 1,239,565 controls). Genome-wide significance is shown for the common threshold of *p*-value < $5x10^{-8}$ (red dashed line). The p-values referenced here correspond to a two-sided Z-test as implemented in METAL.

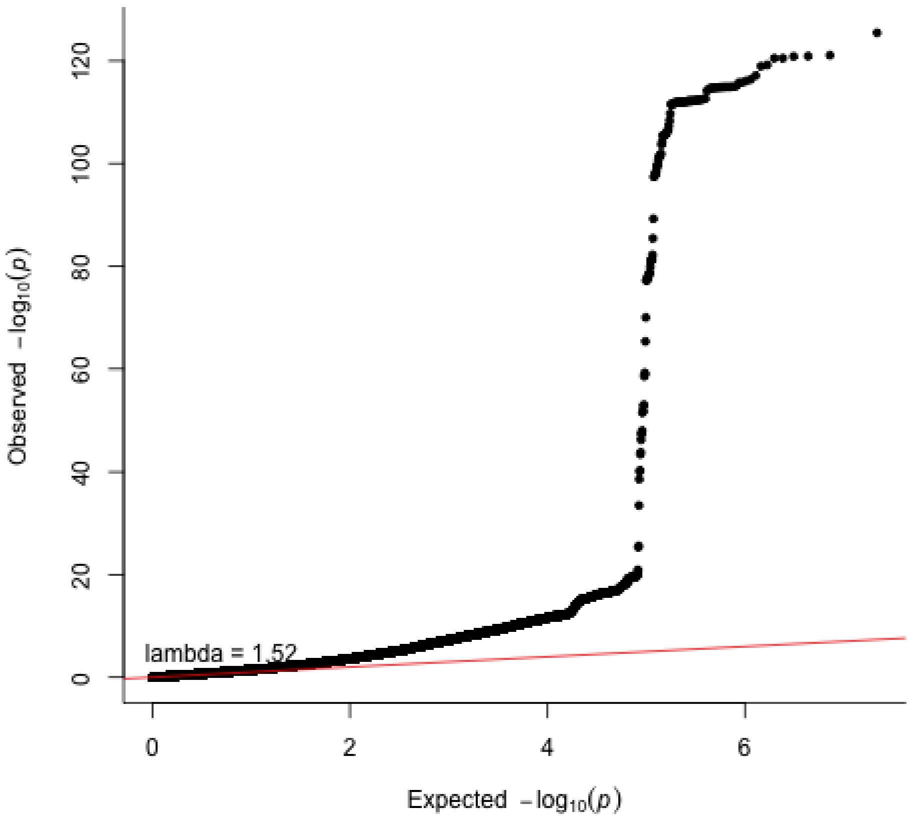


**Supplementary Figure 19. Obstructive sleep apnoea European meta-analysis excluding the Australian cohorts and CLSA QQ plot (n = 1,466,459; 226,894 cases and 1,239,565 controls).** The p-values referenced here correspond to a two-tailed Z-test as implemented in METAL. Each dot compares the observed and expected –log10(P) values. The red diagonal line indicates the expected distribution under the null hypothesis. λGC denotes the genomic inflation factor.

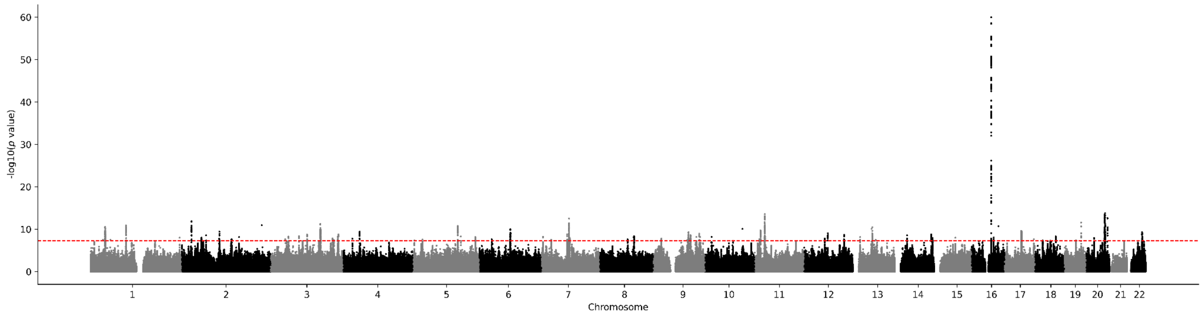


**Supplementary Figure 20. Obstructive sleep apnoea European meta-analysis excluding MVP Manhattan plot.** Results for obstructive sleep apnoea GWAS (n = 1,061,357; 105,597 cases and 955,760 controls). Genome-wide significance is shown for the common threshold of *p*-value < $5x10^{-8}$ (red dashed line). The p-values referenced here correspond to a two-sided Z-test as implemented in METAL.

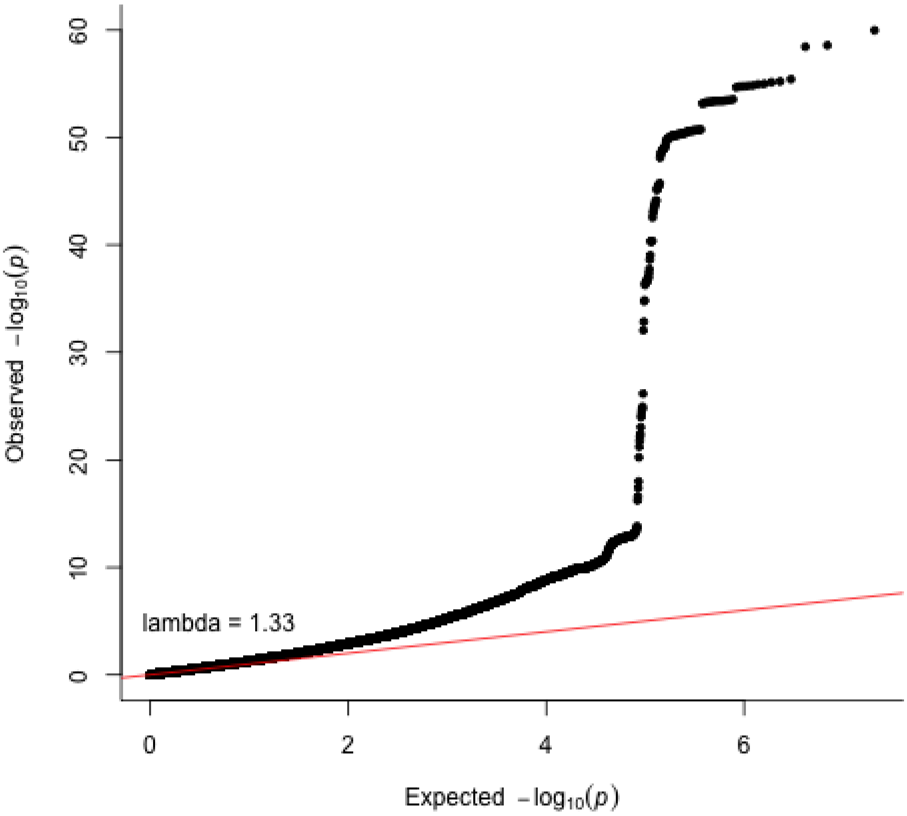


**Supplementary Figure 21. Obstructive sleep apnoea European meta-analysis excluding MVP QQ plot (n = 1,061,357; 105,597 cases and 955,760 controls).** The p-values referenced here correspond to a two-tailed Z-test as implemented in METAL. Each dot compares the observed and expected −log10(P) values. The red diagonal line indicates the expected distribution under the null hypothesis. λGC denotes the genomic inflation factor.

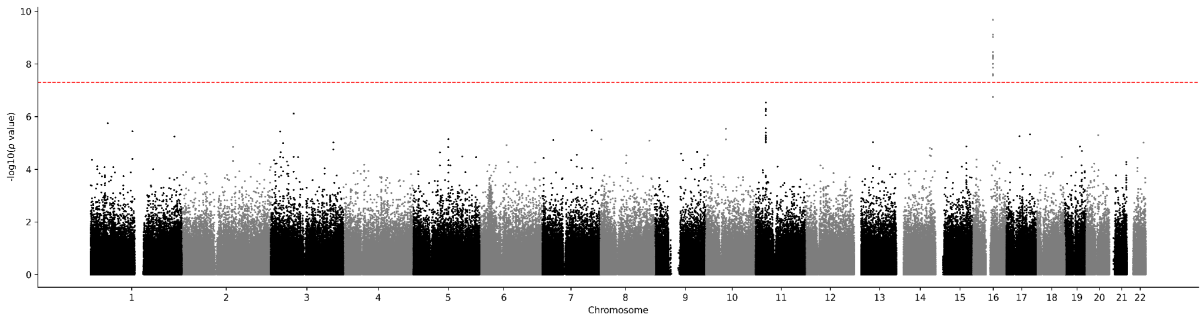


**Supplementary Figure 22. Obstructive sleep apnoea Manhattan plot in UK Biobank sensitivity analysis.** Results for obstructive sleep apnoea GWAS (n = 264,677; 5,959 cases and 258,718 controls). Genome-wide significance is shown for the common threshold of *p*-value < $5x10^{-8}$ (red dashed line). The p-values referenced here correspond to a two-sided score test ($\chi^2{}_1$) as implemented in SAIGE's mixed-model framework.

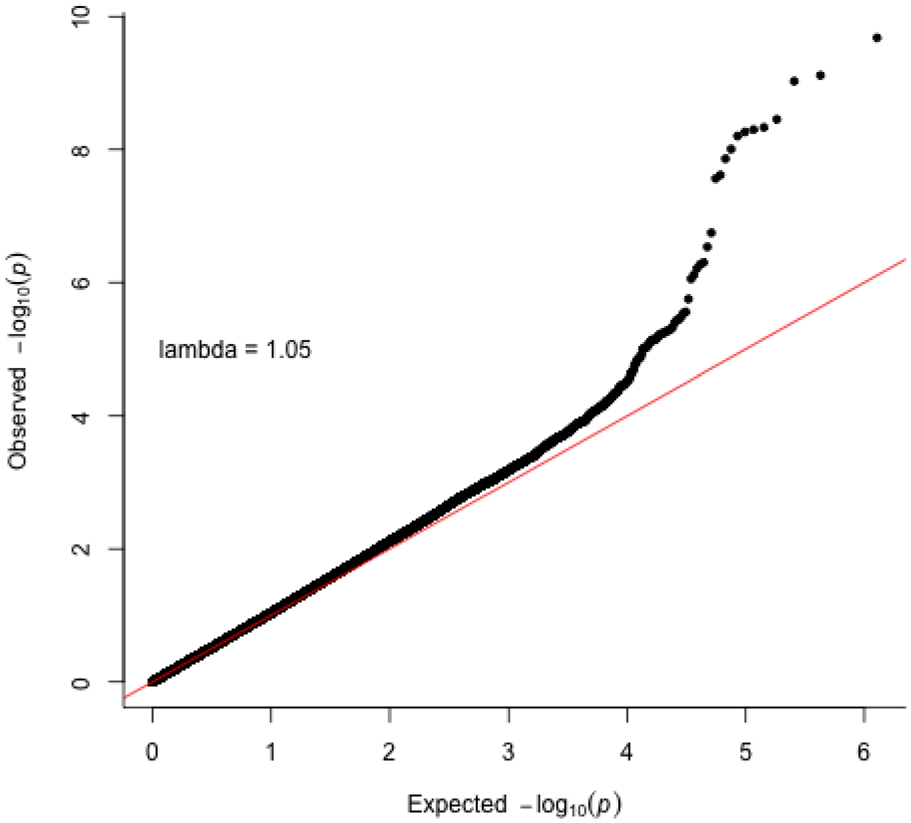


**Supplementary Figure 23. Obstructive sleep apnoea QQ plot in UK Biobank sensitivity analysis (n = 264,677; 5,959 cases and 258,718 controls).** The p-values referenced here correspond to a two-sided score test ($\chi^2{}_1$) as implemented in SAIGE's mixed-model framework. Each dot compares the observed and expected −log10(P) values. The red diagonal line indicates the expected distribution under the null hypothesis. λGC denotes the genomic inflation factor.

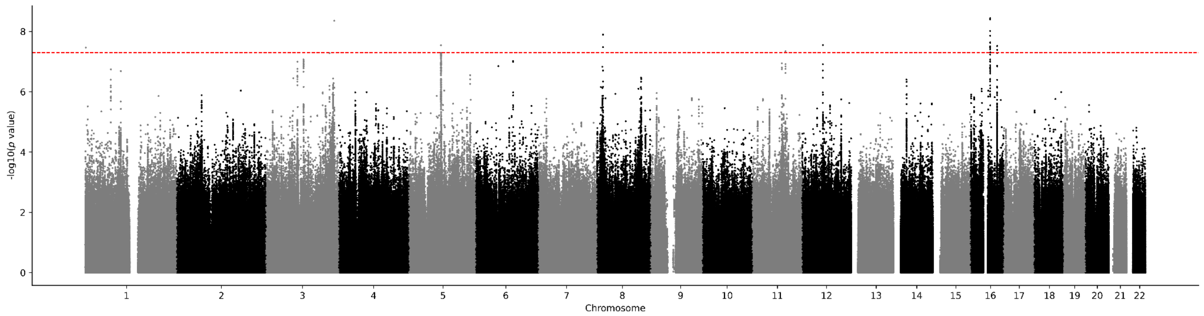


**Supplementary Figure 24. Obstructive sleep apnoea meta-analysis in African ancestry Manhattan plot.** Results for obstructive sleep apnoea GWAS (n = 196,026; 46,834 cases and 149,192 controls). Genome-wide significance is shown for the common threshold of *p*-value < $5x10^{-8}$ (red dashed line). The p-values referenced here correspond to a two-tailed Z-test as implemented in METAL.

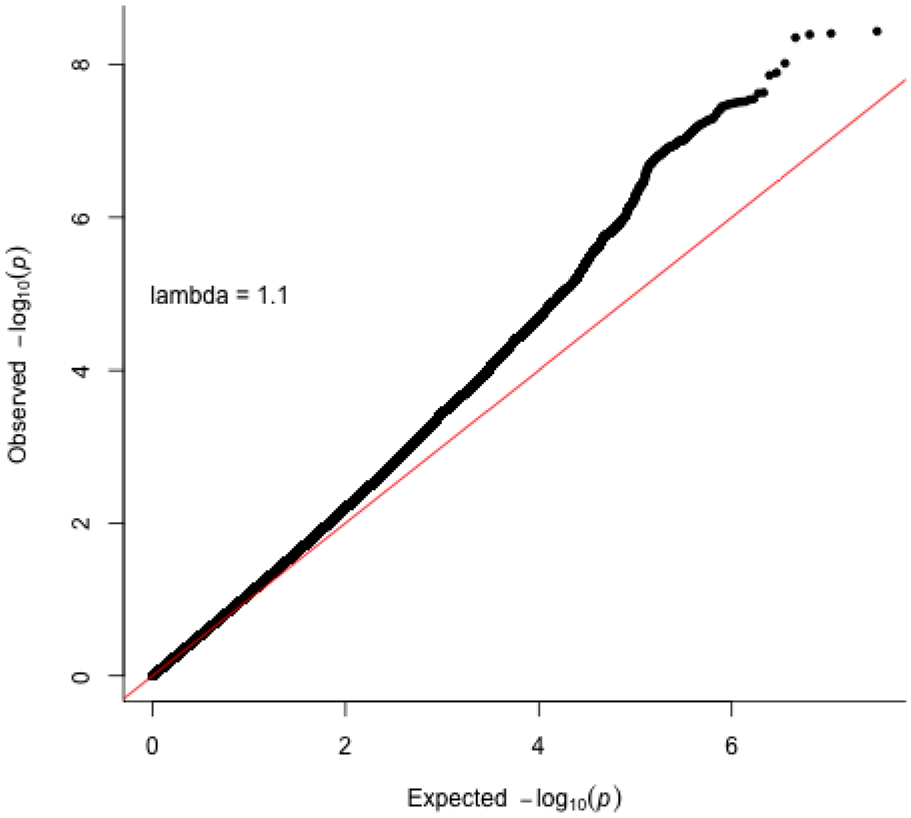


**Supplementary Figure 25. Obstructive sleep apnoea meta-analysis in African ancestry QQ plot (n = 196,026; 46,834 cases and 149,192 controls).** The p-values referenced here correspond to a two-tailed Z-test as implemented in METAL. Each dot compares the observed and expected −log10(P) values. The red diagonal line indicates the expected distribution under the null hypothesis. λGC denotes the genomic inflation factor.

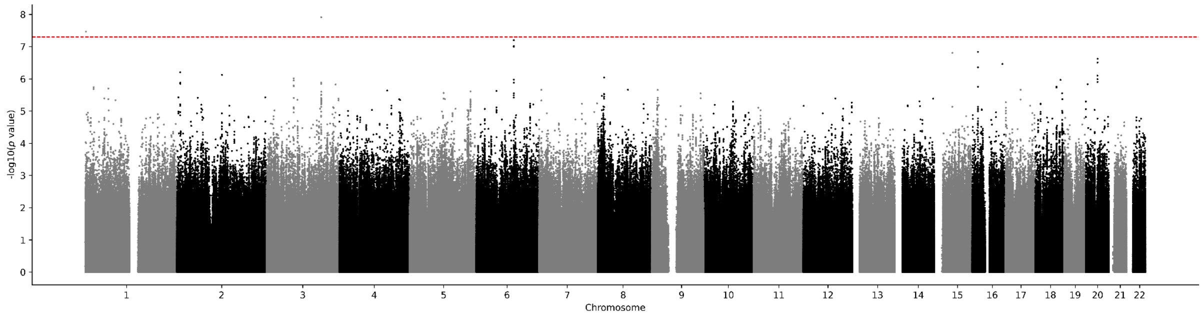


**Supplementary Figure 26. Obstructive sleep apnoea in African ancestry Manhattan plot in AllOfUs.** Results for obstructive sleep apnoea GWAS (n = 80,285; 8,822 cases and 71,463 controls). Genome-wide significance is shown for the common threshold of *p*-value < $5x10^{-8}$ (red dashed line). The p-values referenced here correspond to a two-sided Wald test ($\chi^2_1$) as implemented in REGENIE.

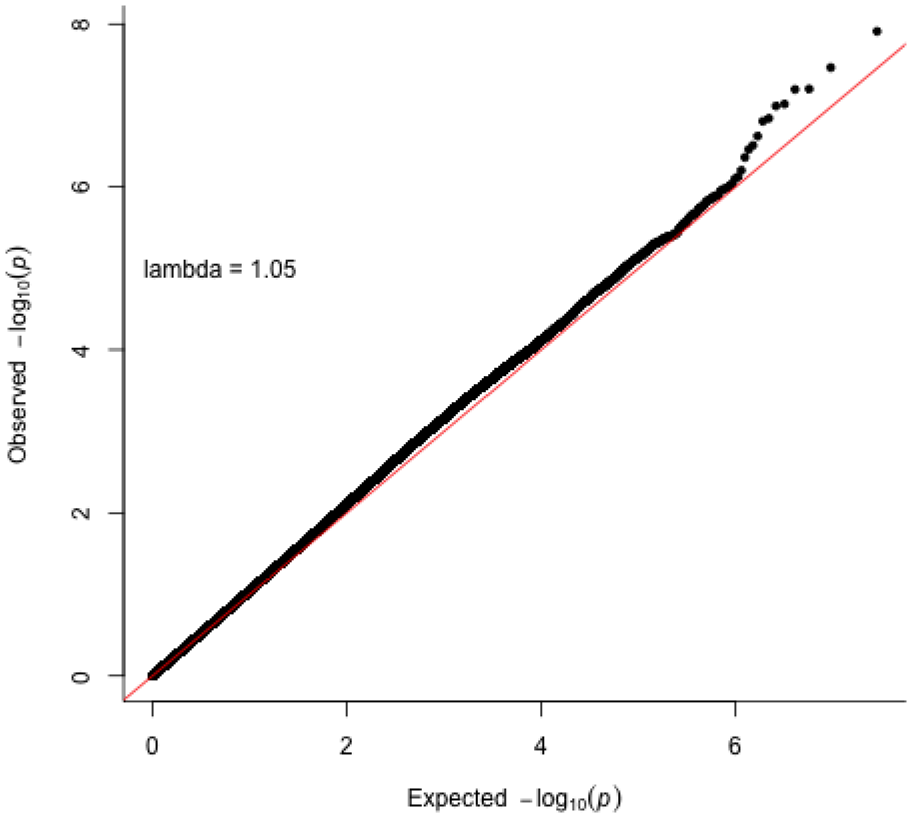


**Supplementary Figure 27. Obstructive sleep apnoea in African ancestry QQ plot in AllOfUs (n = 80,285; 8,822 cases and 71,463 controls).** The p-values referenced here correspond to a two-sided Wald test ($\chi^2_1$) as implemented in REGENIE. Each dot compares the observed and expected −log10(P) values. The red diagonal line indicates the expected distribution under the null hypothesis. λGC denotes the genomic inflation factor.

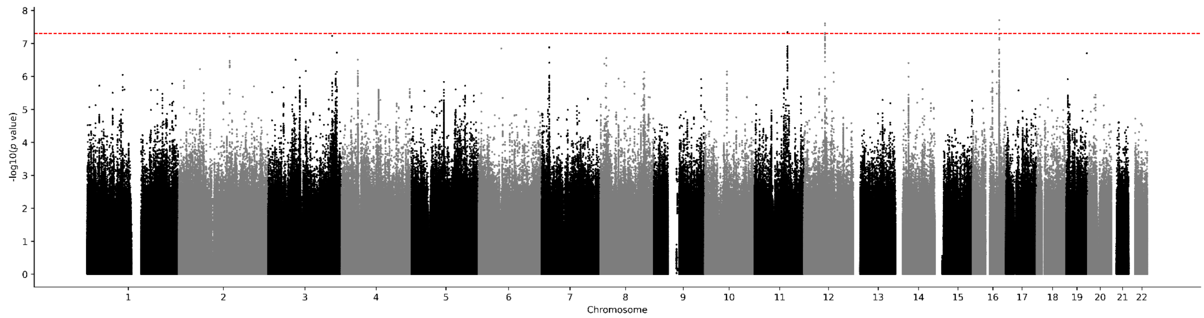


**Supplementary Figure 28. Obstructive sleep apnoea in African ancestry Manhattan plot in MVP.** Results for obstructive sleep apnoea GWAS (n = 115,741; 38,012 cases and 77,729 controls). Genome-wide significance is shown for the common threshold of *p*-value < $5x10^{-8}$ (red dashed line). The p-values referenced here correspond to a two-sided score test ($\chi^2{}_1$) as implemented in SAIGE's mixed-model framework.

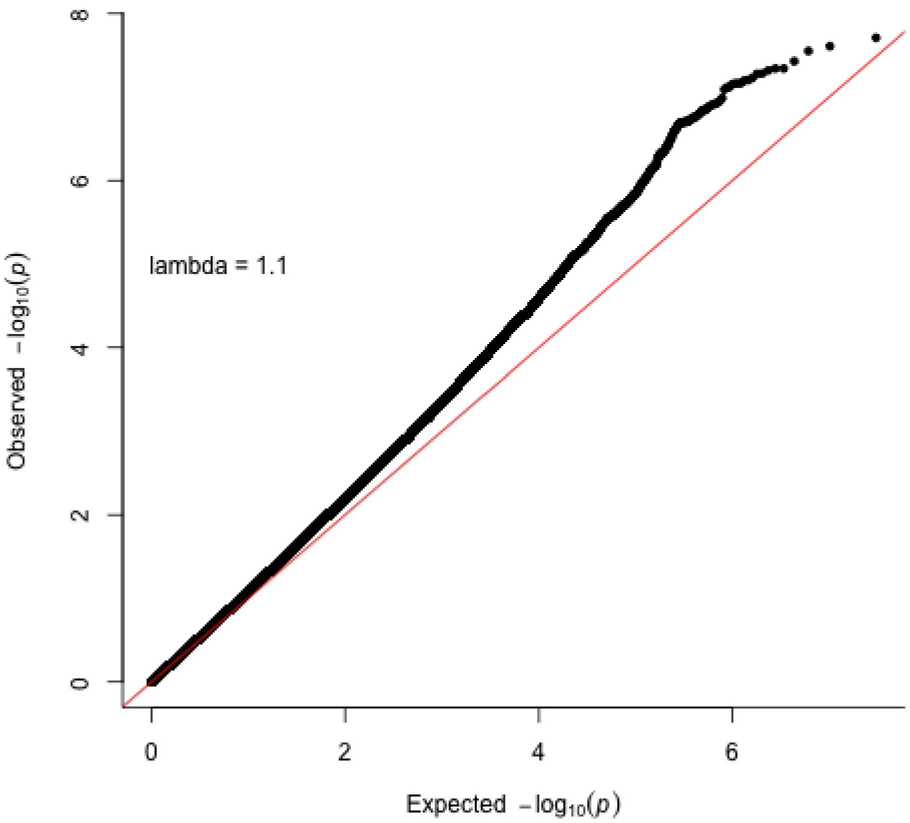


**Supplementary Figure 29. Obstructive sleep apnoea in African ancestry QQ plot in MVP (n = 115,741; 38,012 cases and 77,729 controls).** The p-values referenced here correspond to a two-sided score test ($\chi^2{}_1$) as implemented in SAIGE's mixed-model framework. Each dot compares the observed and expected −log10(P) values. The red diagonal line indicates the expected distribution under the null hypothesis. λGC denotes the genomic inflation factor.

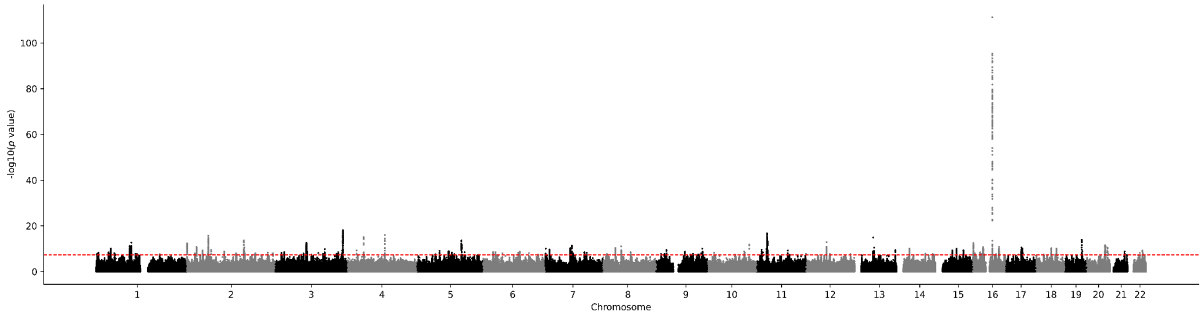


**Supplementary Figure 30. Obstructive sleep apnoea meta-analysis in multi-ancestry Manhattan plot.** Results for obstructive sleep apnoea GWAS (n = 1,751,055; 300,368 cases and 1,450,687 controls). Genome-wide significance is shown for the common threshold of *p*-value < $5x10^{-8}$ (red dashed line). The p-values referenced here correspond to a two-tailed Z-test as implemented in METAL.

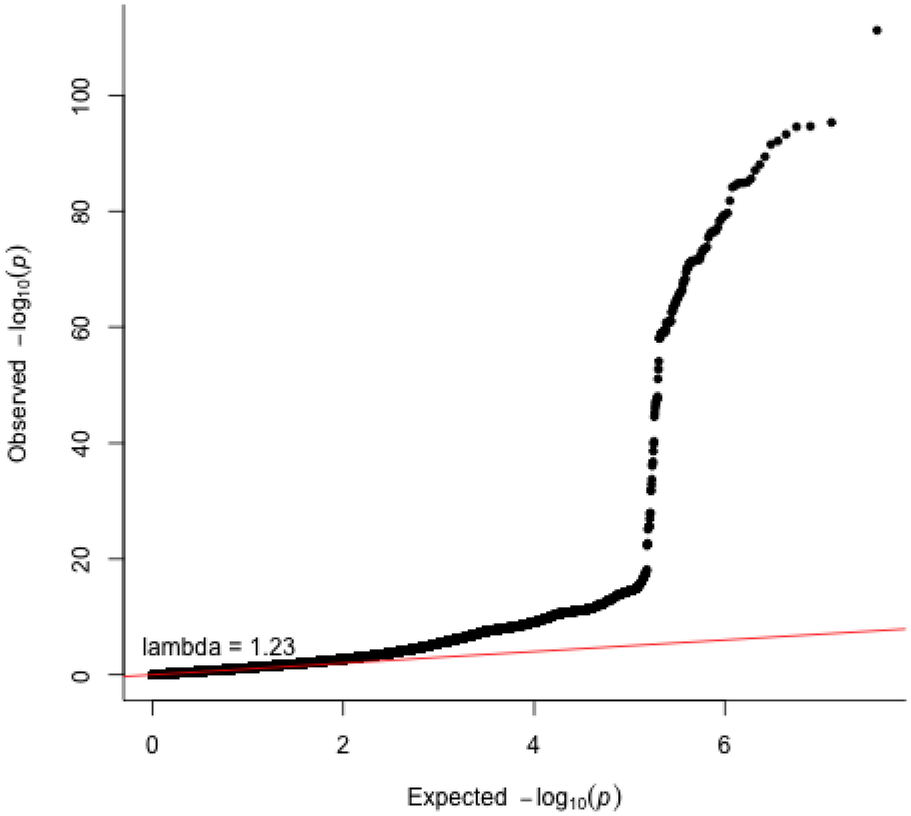


**Supplementary Figure 31. Obstructive sleep apnoea meta-analysis in multi-ancestry ancestry QQ plot (n = 1,751,055; 300,368 cases and 1,450,687 controls).** The p-values referenced here correspond to a two-tailed Z-test as implemented in METAL. Each dot compares the observed and expected −log10(P) values. The red diagonal line indicates the expected distribution under the null hypothesis. λGC denotes the genomic inflation factor.

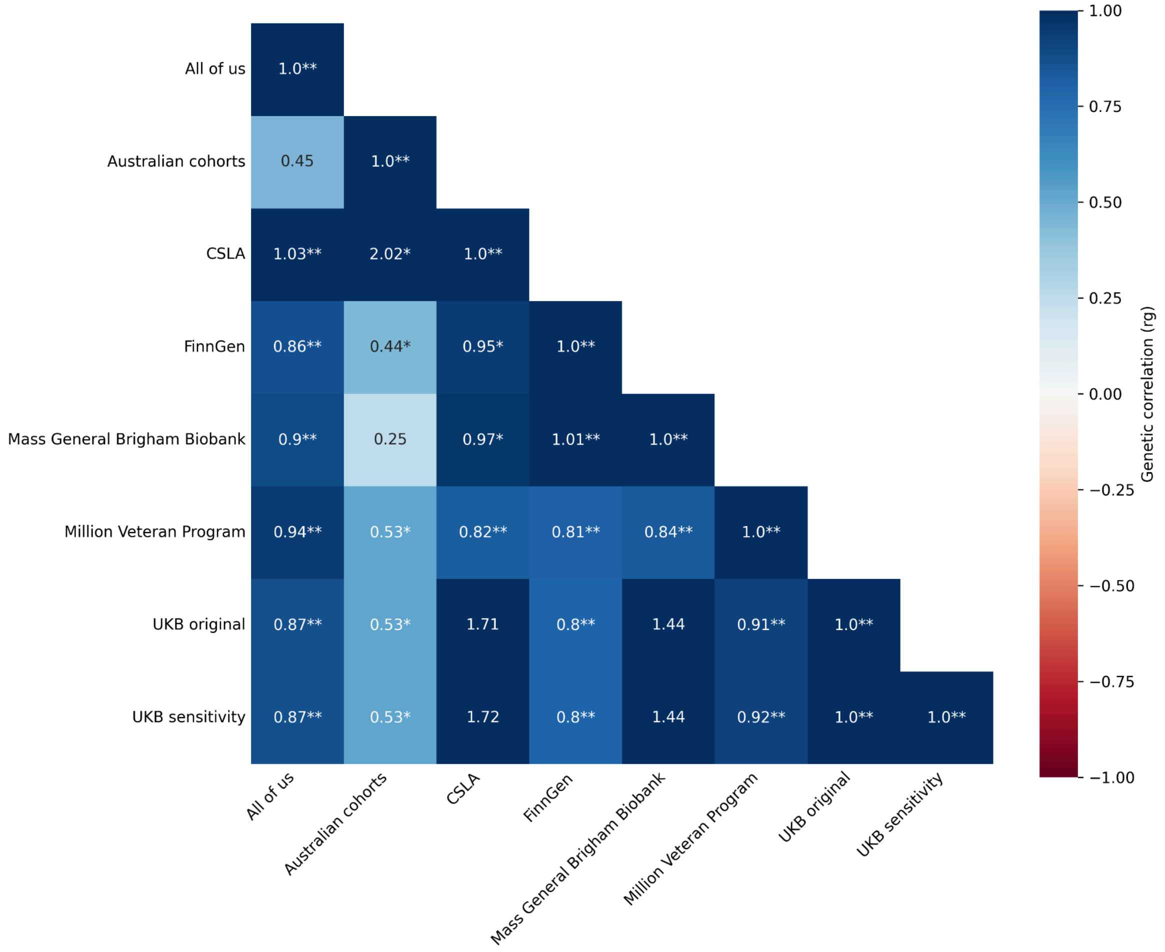


**Supplementary Figure 32. Genetic correlations across European ancestry individual cohorts estimated with LD Score Regression.**

Pairwise genetic correlations were estimated from GWAS summary statistics using bivariate LD Score regression. Values within cells represent genetic-correlation estimates ((r_g)). Blue indicates positive genetic correlation and red indicates negative genetic correlation, with colour intensity representing the magnitude of the estimate. The upper triangle is omitted because the correlation matrix is symmetric, and diagonal values represent self-correlations ((r_g = 1)). LD Score regression estimates are not constrained to the theoretical interval from −1 to 1 and may therefore exceed these bounds owing to sampling uncertainty.

Statistical significance was assessed using a two-sided (z)-test of the null hypothesis (r_g = 0), using the genetic-correlation estimate and its standard error. *(P <) 0.05; **(P <) 0.01.

Australian cohorts comprise the Australian Genetics of Depression Study and the Genetics of Bipolar Disorder project. CLSA, Canadian Longitudinal Study on Aging; MVP, Million Veteran Program; UKB, UK Biobank. “UKB original” denotes the primary UKB OSA GWAS, comprising 5,959 cases and 264,320 controls; participants reporting snoring were excluded from the control group, whereas “UKB sensitivity” represents the sensitivity GWAS using the same 5,959 cases and a more stringent set of 258,718 controls, excluding participants who reported either snoring or frequent daytime sleepiness. Both GWASs used the same analytical framework and variant-level quality-control thresholds. The sensitivity GWAS was conducted to evaluate the influence of possible undiagnosed OSA among controls and was not included in the primary meta-analysis.

### Supplementary references


1. Lavie, L. Oxidative stress in obstructive sleep apnea and intermittent hypoxia--revisited--the bad ugly and good: implications to the heart and brain. *Sleep medicine reviews* **20**, (2015).
2. Pau, M. C. *et al.* Evaluation of Inflammation and Oxidative Stress Markers in Patients with Obstructive Sleep Apnea (OSA). *Journal of Clinical Medicine* **12**, 3935 (2023).
3. Huang, Y. & Mahley, R. W. Apolipoprotein E: Structure and Function in Lipid Metabolism, Neurobiology, and Alzheimer's Diseases. *Neurobiology of disease* **72PA**, 3 (2014).
4. Chamberlin, N. L. Brain circuitry mediating arousal from obstructive sleep apnea. *Current opinion in neurobiology* **23**, (2013).
5. Tran, L. T. *et al.* Hypothalamic control of energy expenditure and thermogenesis. *Experimental & Molecular Medicine* **54**, 358–369 (2022).
6. Bulik-Sullivan, B. K. *et al.* LD Score regression distinguishes confounding from polygenicity in genome-wide association studies. *Nature genetics* **47**, (2015).
7. Benjafield, A. V. *et al.* Estimation of the global prevalence and burden of obstructive sleep apnoea: a literature-based analysis. *The Lancet. Respiratory medicine* **7**, 687 (2019).
8. Malhotra, A. *et al.* Metrics of sleep apnea severity: beyond the apnea-hypopnea index. *Sleep* **44**, zsab030 (2021).
9. Rana, A. M. & Sankari, A. Central Sleep Apnea. in *StatPearls [Internet]* (StatPearls Publishing, 2023).

10. Sofer, T. *et al.* Genome-wide association study of obstructive sleep apnoea in the Million Veteran Program uncovers genetic heterogeneity by sex. *EBioMedicine* **90**, 104536 (2023).
11. Genomic data in the All of Us Research Program. *Nature* **627**, (2024).
12. Vuokko, R., Vakkuri, A. & Palojoki, S. Systematized Nomenclature of Medicine–Clinical Terminology (SNOMED CT) Clinical Use Cases in the Context of Electronic Health Record Systems: Systematic Literature Review. *JMIR Medical Informatics* **11**, e43750 (2023).
13. Mbatchou, J. *et al.* Computationally efficient whole-genome regression for quantitative and binary traits. *Nature Genetics* **53**, 1097–1103 (2021).
14. Lind, P. A. *et al.* Preliminary results from the Australian Genetics of Bipolar Disorder Study: A nation-wide cohort. *The Australian and New Zealand Journal of Psychiatry* **57**, 1428 (2023).
15. Byrne, E. M. *et al.* Cohort profile: the Australian genetics of depression study. *BMJ Open* **10**, e032580 (2020).
16. Das, S. *et al.* Next-generation genotype imputation service and methods. *Nat. Genet.* **48**, 1284–1287 (2016).
17. Zhou, W. *et al.* Efficiently controlling for case-control imbalance and sample relatedness in large-scale genetic association studies. *Nature Genetics* **50**, 1335–1341 (2018).
18. Raina, P. *et al.* Cohort Profile: The Canadian Longitudinal Study on Aging (CLSA). *International Journal of Epidemiology* **48**, 1752 (2019).
19. Raina, P. S. *et al.* The Canadian longitudinal study on aging (CLSA). *Canadian journal on aging = La revue canadienne du vieillissement* **28**, (2009).

20. Forgetta, V. *et al.* Cohort profile: genomic data for 26 622 individuals from the Canadian Longitudinal Study on Aging (CLSA). *BMJ Open* **12**, e059021 (2022).

21. Sudlow, C. *et al.* UK biobank: an open access resource for identifying the causes of a wide range of complex diseases of middle and old age. *PLoS Med.* **12**, e1001779 (2015).

22. Bycroft, C. *et al.* The UK Biobank resource with deep phenotyping and genomic data. *Nature* **562**, 203–209 (2018).

23. Campos, A. I. *et al.* Discovery of genomic loci associated with sleep apnea risk through multi-trait GWAS analysis with snoring. *Sleep* **46**, (2023).

24. Campos, A. I. *et al.* Insights into the aetiology of snoring from observational and genetic investigations in the UK Biobank. *Nature Communications* **11**, 1–12 (2020).

25. Kurki, M. I. *et al.* FinnGen provides genetic insights from a well-phenotyped isolated population. *Nature* **613**, (2023).

26. Verma, A. *et al.* Diversity and scale: Genetic architecture of 2068 traits in the VA Million Veteran Program. *Science* https://doi.org/10.1126/science.adj1182 (2024) doi:10.1126/science.adj1182.

27. Karlson, E. W., Boutin, N. T., Hoffnagle, A. G. & Allen, N. L. Building the Partners HealthCare Biobank at Partners Personalized Medicine: Informed Consent, Return of Research Results, Recruitment Lessons and Operational Considerations. *Journal of personalized medicine* **6**, (2016).

28. Boutin, N. T. *et al.* The Evolution of a Large Biobank at Mass General Brigham. *Journal of Personalized Medicine* **12**, 1323 (2022).

29. Purcell, S. *et al.* PLINK: A Tool Set for Whole-Genome Association and Population-Based Linkage Analyses. *American Journal of Human Genetics* **81**, 559 (2007).

30. de Leeuw, C. A., Mooij, J. M. & Heskes, T. MAGMA: generalized gene-set analysis of GWAS data. *PLoS computational biology* **11**, (2015).

31. Watanabe, K., Taskesen, E. & van Bochoven, A. Functional mapping and annotation of genetic associations with FUMA. *Nature communications* **8**, (2017).

32. Bakshi, A. *et al.* Fast set-based association analysis using summary data from GWAS identifies novel gene loci for human complex traits. *Scientific Reports* **6**, 1–9 (2016).

33. Lonsdale, J. *et al.* The Genotype-Tissue Expression (GTEx) project. *Nature Genetics* **45**, 580–585 (2013).

34. Finucane, H. K. *et al.* Heritability enrichment of specifically expressed genes identifies disease-relevant tissues and cell types. *Nature genetics* **50**, 621 (2018).

35. Mancuso, N. *et al.* Probabilistic fine-mapping of transcriptome-wide association studies. *Nature genetics* **51**, 675 (2019).

36. Zhu, Z. *et al.* Integration of summary data from GWAS and eQTL studies predicts complex trait gene targets. *Nature Genetics* **48**, 481–487 (2016).

37. García-Marín, L. M. *et al.* Genomic analysis of intracranial and subcortical brain volumes yields polygenic scores accounting for variation across ancestries. *Nature Genetics* **56**, 2333–2344 (2024).

38. Wu, K. *et al.* Obstructive sleep apnea and structural and functional brain alterations: a brain-wide investigation from clinical association to genetic causality. *BMC Medicine* **23**, 1–11 (2025).

39. Joo, E. Y., Jeon, S., Kim, S. T., Lee, J.-M. & Hong, S. B. Localized Cortical Thinning in Patients with Obstructive Sleep Apnea Syndrome. *Sleep* **36**, 1153 (2013).

40. Structural and functional neural adaptations in obstructive sleep apnea: An activation likelihood estimation meta-analysis. *Neuroscience & Biobehavioral Reviews* **65**, 142–156 (2016).

41. Grasby, K. L. *et al.* The genetic architecture of the human cerebral cortex. *Science* https://doi.org/10.1126/science.aay6690 (2020) doi:10.1126/science.aay6690.

42. Sullivan, P. F. *et al.* Psychiatric Genomics: An Update and an Agenda. *The American journal of psychiatry* **175**, (2018).

43. Middeldorp, C. M., Felix, J. F., Mahajan, A. & McCarthy, M. I. The Early Growth Genetics (EGG) and EArly Genetics and Lifecourse Epidemiology (EAGLE) consortia: design, results and future prospects. *European journal of epidemiology* **34**, (2019).

44. Pickrell, J. K. *et al.* Detection and interpretation of shared genetic influences on 42 human traits. *Nature genetics* **48**, (2016).

45. García-Marín, L. M. *et al.* Shared molecular genetic factors influence subcortical brain morphometry and Parkinson's disease risk. *npj Parkinson's Disease* **9**, 1–10 (2023).

46. Reyes-Pérez, P. *et al.* Investigating the Shared Genetic Etiology Between Parkinson's Disease and Depression. *Journal of Parkinson's Disease* https://doi.org/10.3233/JPD-230176 (2024) doi:10.3233/JPD-230176.

47. García-Marín, L. M. *et al.* Investigating the genetic relationship of intracranial and subcortical brain volumes with depression and other psychiatric disorders. *Imaging Neuroscience* https://doi.org/10.1162/imag_a_00291 (2024) doi:10.1162/imag_a_00291.

48. Burgess, S., Foley, C. N., Allara, E., Staley, J. R. & Howson, J. M. M. A robust and efficient method for Mendelian randomization with hundreds of genetic variants. *Nature Communications* **11**, 1–11 (2020).

49. Darrous, L., Mounier, N. & Kutalik, Z. Simultaneous estimation of bi-directional causal effects and heritable confounding from GWAS summary statistics. *Nature communications* **12**, (2021).

### Supplementary methods

#### Cohorts and GWAS for European populations

***All of Us.*** The All of Us research program is a longitudinal cohort study seeking to recruit at least one million individuals from diverse ancestral backgrounds across the USA by 2027[11]. We performed GWAS for OSA (32,618 cases and 192,980 controls) using whole-genome sequencing data for participants of European ancestry from All of Us version 8. The whole genome sequencing data were processed using the Illumina DRAGEN pipeline (v3.7.8)[11]. Ancestry was defined using the genetically inferred ancestry provided by the All of Us research program, computed through Rye[11]. OSA was defined based on the (Systematised Nomenclature of Medicine) SNOMED[12] vocabulary concept ID 78275009. Participants who self-reported snoring were removed from the controls in the OSA GWAS due to the possibility of representing undiagnosed OSA. GWAS were conducted using REGENIE[13] including age, sex, and 10 principal components as covariates. During quality control, we excluded variants with a low minor allele count (<20) and minor allele frequency (<0.01) from the analysis.

***Australian cohorts.*** We leveraged data from participants of European ancestry from the Australian Genetics of Bipolar Disorder Study (GBP)[14] and the Australian Genetics of Depression Study (AGDS)[15] to conduct GWAS for OSA. A thorough description for each of these cohorts is available elsewhere[14,15]. Briefly, GBPis a nationwide cohort of adults living with bipolar disorder. So far, GBP includes ~7,000 participants, of which 70% have been genotyped[14]. Similarly, AGDS is a nation-wide cohort study of individuals who have been diagnosed with depression at some point in their lifetime[15]. AGDS includes ~20,000 participants, of which ~18,000 have been genotyped[15]. Both cohort studies were genotyped with the Illumina Global Screening Array v2, leveraged the Michigan imputation server with the Haplotype Reference Consortium reference panel[16] to impute the genotypes and performed similar cohort quality control procedures[14,15]. In addition, both studies included the same items in their self-reported questionnaires to assess OSA. Therefore, we were able to conduct GWAS for OSA using the samples from both cohorts as a larger single sample.

In the two Australian cohorts, OSA was defined using two items: *'During the last month, on how many nights or days per week have you had or been told you had snorting or gasping?'* and '*During the last month, on how many nights or days per week have you had or been told* *you had your breathing stop or you choked or struggled for breath?'* Participants were classified as OSA cases (N = 1,247) if they reported a frequency anywhere between 1 to 7 times per week for either item. In addition to this, for an individual to be considered a case for OSA, the individual must have been deemed as a snorer. These items capture symptom domains overlapping conceptually with validated screening instruments for OSA (e.g., observed apnoea components of STOP-Bang), but the cohorts did not administer the full STOP-Bang questionnaire or an equivalent validated screening tool. We note that these questionnaire items are not part of a formally validated OSA screening instrument and were not directly validated against clinical diagnoses such as polysomnography. Accordingly, these definitions reflect symptom-based self-report consistent with probable or possible OSA rather than clinically confirmed disease. The OSA control group (N = 8,389) included individuals who did not snore (*Rarely, less than once a week* or *Never*) or have OSA. The OSA control group also excluded those with snoring but no OSA. GWAS for OSA was performed using SAIGE[17] and a generalised linear mixed model. The GWAS was adjusted for age and sex, and 10 principal components. We excluded variants with a low minor allele frequency (<0.01) or a low-quality imputation score (<0.60) from the analysis.

***Canadian Longitudinal Study on Aging (CLSA).*** The Canadian Longitudinal Study of Ageing (CLSA) seeks to understand and address the needs of an ageing population. It includes ~50,000 participants AGED 45 to 85 years at the time of recruitment, of which ~20,000 have been genotyped via Affymetrix Axiom array and imputed through the Michigan Imputation Server using the TOPMed reference panel[18]. A thorough description of this cohort is available elsewhere[18–20]. In this cohort, OSA prevalence (4,151 cases and 11,436 controls) was defined via the categories "SNO_STOPBREATH_MCQ" and "SNO_STOPBREATH_COF1" from baseline and follow-up surveys, respectively. These categories include the item *'stop breathing in sleep',* with '*Yes*', '*No*', and *'Don't know'* as possible responses. At baseline, 14.7% responded '*Yes*', while 83.2% responded *'No'*. In the follow-up, 16.8% responded '*Yes*', and 78.3% '*No*'. In this analysis, we excluded participants who responded '*Don't know*'. In addition, if a participant reported *'Yes'* at baseline but 'No' in the follow-up survey or vice versa, they were deemed as cases since they reported OSA at least once. Participants who self-reported snoring were also removed from the controls in the OSA GWAS due to the possibility of representing undiagnosed OSA. As in the Australian cohorts, we note that these questionnaire items are not part of a formally validated OSA screening instrument and were not directly validated against clinical diagnoses such as polysomnography. Accordingly, these definitions reflect symptom-based self-report consistent with probable or possible OSA rather than clinically confirmed disease. The GWAS included individuals of European ancestry for OSA and was conducted using REGENIE(v2.2.4)[13], including age, sex, and 10 principal components as covariates. We excluded variants with a low minor allele frequency (<0.01) or a low-quality imputation score (<0.60) from the analysis. Also, the Regenie '--firth' flag was used to perform Firth correction to provide a reduction in bias due to the relatively small sample size[13].

***UK Biobank (UKB).*** We conducted GWAS for OSA (5,959 cases and 264,320 controls) using data for participants of European ancestry from the UK Biobank[21]. Genotyping details, which included using the UK BiLEVE Axiom Array for the first ~50,000 participants and the UK Biobank Axiom Array for the remaining ~450,000, as well as phenotyping details for this cohort have been described elsewhere[22]. Briefly, for consistence with previous studies[23,24], we defined OSA through the ICD-10 sleep apnoea diagnosis code (code G47.3 in the 41270 UKB data field), and self-reported sleep apnoea (code 1123 in the 20002 UKB data field). Participants with only *'Yes'* or *'No'* responses were included, leaving out those who answered '*I don't know*' or *'Prefer not to answer'.* Participants who self-reported snoring (defined using data field 1210) were removed from the controls in the OSA GWAS due to the possibility of representing undiagnosed OSA. We conducted the GWAS using SAIGE[17] and a mixed generalised linear model, and included age, sex, genotyping array, and 10 principal components as covariates to adjust further for population stratification. We excluded variants with a low minor allele frequency (<0.01) or a low-quality imputation score (<0.60) from the analysis.

***UKB sensitivity analysis.*** To assess the robustness of the UKB OSA GWAS to potential misclassification of undiagnosed OSA among controls, we conducted a sensitivity analysis using a more stringent control definition. In this analysis, controls were restricted to participants without self-reported snoring (UKB data field 1210) and/or without frequent daytime sleepiness (UKB data field 1220). Daytime sleepiness was defined via the question *'How likely are you to doze off or fall asleep during the daytime when you don't mean to? (e.g. when working, reading or driving)'* (UK Biobank data field 1220). Individuals who responded *'Often'* or *'All the time'*, were considered daytime sleepiness cases, and those who responded *'Do not know'* or *'Prefer not to answer'* were excluded.

Under this stricter control definition, the sensitivity UKB GWAS included 5,959 OSA cases and 258,718 controls. The GWAS was performed using the same analytical framework as the primary UKB analysis, including SAIGE with a mixed generalised linear model and adjustment for age, sex, genotyping array, and the first 10 principal components of ancestry. Variant-level quality control thresholds were identical to those applied in the primary analysis (minor allele frequency < 0.01 and imputation quality < 0.60 excluded). This sensitivity analysis was conducted to evaluate the impact of alternative control definitions on association results and was not used as input to the primary meta-analysis.

***FinnGen.*** We leveraged publicly available GWAS summary statistics for obstructive sleep apnoea (OSA) from the FinnGen cohort Data Freeze 12 (R12), comprising participants of European ancestry[25]. A detailed description of the FinnGen study design, genotyping, and phenotyping procedures has been published previously and is provided by the FinnGen consortium and is available elsewhere[25]. OSA was defined using the ICD-10 diagnosis code G47.3 (sleep apnoea), corresponding to the FinnGen endpoint G6_SLEEPAPNO[25]. GWAS summary statistics were generated using a linear mixed model implemented in SAIGE, with adjustment for sex, age, genotyping batch, and the first 10 principal components of ancestry[25]. The analysis included a total of 58,520 cases and 461,690 controls[25]. Variants were filtered to retain those with imputation quality INFO > 0.60 and minor allele frequency > 0.01.

***Million Veteran Program (MVP).*** We obtained publicly available GWAS summary statistics, including participants of European ancestry for OSA from MVP, genotyped via the MVP 1.0 Axiom array and imputed using the TOPMed reference panel. The summary data included 126,695 cases and 303,630 controls and were derived as part of the MVP genome-wide PheWAS project[26] in a collaboration between the US Department of Veterans Affairs and the US Department of Energy[26]. OSA was defined via electronic health records using a multimodal automated phenotyping procedure, which analyses ICD codes and natural language processing of clinical notes. GWAS were conducted using a linear mixed model approach in SAIGE[17] adjusting for age, sex, and the first 10 principal components[26]. Further details regarding GWAS summary statistics from MVP are available elsewhere[26]. We performed quality control procedures on the GWAS summary data and excluded variants with a low minor allele frequency (<0.01) or a low-quality imputation score (<0.60).

***Mass General Brigham Biobank.*** We leveraged GWAS summary statistics for OSA (3,102 cases and 16,945 controls)[23], including participants of European ancestry from the Mass General Brigham Biobank (formerly the Partners Healthcare Biobank)[27,28], generated in a previous study[23]. Full details about the Mass General Brigham Biobank are available elsewhere[27,28]. Briefly, this Biobank focuses on electronic health records and survey data, including ~135,000 participants, for which ~65,000 have genomic data available[27,28]. Data was genotyped using the Illumina Multi-Ethnic Global Array (MEGA) (Illumina, Inc., San Diego, CA) GWAS array[27,28], and imputation was performed on the Michigan Imputation server[23]. OSA was defined using electronic health records data and the ICD-10 sleep apnoea diagnosis code (G47.3)[23]. The GWAS was conducted using PLINK[29] v2.00 and a logistic regression with age, sex, genetic principal components, and genotype batch as covariates[23]. The GWAS excluded variants with a low minor allele frequency (<0.01)[23].

**GWAS meta-analysis in European populations**

***METAL.*** Genomic control was not applied at the meta-analysis stage. The genomic inflation factor (λGC) was calculated from the GWAS test statistics as the ratio between the observed median $\chi^2$ statistic and the expected median of the null $\chi^2$ distribution with one degree of freedom (0.455). The genomic inflation factor for the meta-analysis was λGC = 1.51 (**Supplementary Figure 1**). Because λGC increases with sample size and trait polygenicity, we used LD Score regression to evaluate whether the observed inflation reflected residual confounding or true polygenic signal. The LD Score regression intercept was close to unity (intercept = 1.005, SE = 0.0093), with an attenuation ratio of 0.009, indicating that the majority of inflation was attributable to polygenic signal rather than uncorrected population stratification. Accordingly, post-hoc genomic control was not applied, as it can be overly conservative in large, well-powered GWAS and may attenuate true associations.

***Sensitivity analyses.*** To assess the robustness of the meta-analysis to potential phenotype heterogeneity and cohort-specific effects, we performed two complementary sensitivity meta-analyses. First, to evaluate the impact of symptom-based self-report phenotype definitions, we repeated the meta-analysis excluding the Australian cohorts and the Canadian Longitudinal Study on Aging (CLSA). Second, to assess whether results were driven by a single large contributing cohort, we conducted a leave-one-out meta-analysis excluding the Million Veteran Program (MVP), the largest individual cohort. Meta-analysis procedures, quality-control thresholds, and parameters were identical to those used in the primary meta-analysis. Effect estimates and association statistics from both sensitivity analyses are reported in **Supplementary Table 1** and were compared directly with results from the full meta-analysis to assess consistency in effect size magnitude, direction, and statistical significance. For each variant, heterogeneity metrics including the Q statistic, $I^2$, and heterogeneity P-value were computed and are reported in **Supplementary Table 1**.

**Conditional analysis**

***Body mass index genetic effect attenuation.*** To evaluate the locus-level behaviour of BMI conditioning and assess potential collider-like artefacts, we performed two complementary supplementary analyses. First, for each lead variant from the primary OSA meta-analysis, we quantified the proportional attenuation of effect size following BMI conditioning using the attenuation ratio (defined as $OSA_{mtCOJO\_BMI}$ β divided by OSA β, as well as the absolute change in effect size ($\Delta\beta = OSA_{mtCOJO\_BMI}\ \beta - OSA\ \beta$), and compared these metrics with the corresponding effect estimates and p-values from an independent BMI GWAS (**Supplementary Table 1**). All alleles were harmonised to the OSA meta-analysis effect allele prior to comparison. Second, we conducted a targeted directionality screen across the 39 independent genome-wide significant $OSA_{mtCOJO\_BMI}$ loci by comparing aligned effect directions across OSA, $OSA_{mtCOJO\_BMI}$, and BMI GWAS summary statistics (**Supplementary Table 5**). For each locus, we recorded whether all three GWAS datasets shared the same effect size direction, whether a directionality flip occurred between OSA and $OSA_{mtCOJO\_BMI}$, and whether loci satisfied conservative collider patterns at genome-wide BMI loci ($P < 5 \times 10^{-8}$), defined as either BMI $\beta < 0$ with $OSA_{mtCOJO\_BMI}$ $\beta > 0$ or BMI $\beta > 0$ with $OSA_{mtCOJO\_BMI}$ $\beta < 0$, with stricter flags additionally requiring a directionality flip between OSA and $OSA_{mtCOJO\_BMI}$. These annotations were used as sensitivity screens to identify loci potentially sensitive to BMI conditioning, rather than to assert definitive collider bias or BMI independence at individual loci.

**Cohorts and GWAS for African populations**

***All of Us.*** We performed GWAS for OSA (8,822 cases and 71,463 controls) using whole genome sequencing data for participants of African ancestry from All of Us version 8. OSA was defined based on the SNOMED[12] vocabulary concept ID 78275009. Participants who self-reported snoring were removed from the controls in the OSA GWAS due to the possibility of representing undiagnosed OSA. GWAS were conducted using REGENIE(v3.2.6),[13] including age, sex, and 10 principal components as covariates. During quality control, we excluded variants with a low minor allele count (<20) and minor allele frequency (<0.05) from the analysis.

***Million Veteran Program (MVP).*** We obtained publicly available GWAS summary statistics, including participants of African ancestry for OSA from MVP. The summary data included 38,012 cases and 77,729 controls. GWAS were conducted using a linear mixed model approach in SAIGE[17] adjusting for age, sex, and the first 10 principal components[26]. Further details regarding GWAS summary statistics from MVP are available elsewhere[26]. We performed quality control procedures on the GWAS summary data and excluded variants with a low minor allele frequency (<0.01) or a low-quality imputation score (<0.60).

**Functional annotation and gene prioritisation**

We conducted MAGMA[30] gene-based and gene-set (v1.08) analyses as implemented in FUMA (v1.7.0)[31] (https://fuma.ctglab.nl/). Briefly, MAGMA aggregates association *P* values considering all the genetic variants mapping to a given gene and its corresponding regulatory region[30]. We applied a Bonferroni multiple-testing correction (0.05/18,871 (total number of tested genes) = $2.65 \times 10^{-06}$). As a complementary approach to MAGMA's gene-based test, we performed fastBAT (v1.94.1)[32] as part of our initial gene discovery phase and applied a Bonferroni multiple-testing correction (0.05/24,568 (total number of tested genes) = $2.04 \times 10^{-06}$). MAGMA gene-set tests assess predefined biological pathways and GO terms for enrichment of OSA-associated genes, highlighting potential dysregulated processes.

We used MAGMA's Tissue Expression Analysis using GTEx v8[33] as implemented in FUMA to identify tissue specificity of genes associated with OSA. We implemented a Bonferroni multiple-testing correction for general (0.05/30 (total number of general tissues) = $1.67 \times 10^{-03}$) and specific tissues (0.05/54 (total number of specific tissues) = $9.26 \times 10^{-04}$). Furthermore, we investigated tissue-specific enrichment (for those identified with MAGMA's Tissue Expression Analysis) with LDSC-SEG (v1.0.1)[34], a method that integrates GWAS summary data with gene expression patterns via stratified LD score regression to identify tissues or cell types where OSA heritability is enriched. We implemented a Bonferroni multiple-testing correction (0.05/24 (total number of tested tissues) = $2.08 \times 10^{-03}$). In addition, we performed eQTL mapping in FUMA to map OSA-associated SNPs to potential target genes via the expression quantitative trait loci based on 13 GTEx (v8) brain tissues. We applied Bonferroni multiple testing correction and accounted for the number of brain tissues in our analysis (0.05 / 15,757 [average number of annotations per brain volume] * 13 [number of tested brain tissues] = $2.44 \times 10^{-07}$). We performed transcriptome-wide association study (TWAS)-based fine-mapping using FOCUS (Fine-mapping Of CaUsal gene Sets; v0.6)[35] to prioritise candidate causal genes underlying GWAS signals. Gene expression prediction models were derived from GTEx v8 across 13 brain tissues. For each genomic region, FOCUS integrates TWAS summary statistics with linkage disequilibrium (LD) information to estimate posterior inclusion probabilities (PIP) for each gene, representing the probability that predicted gene expression mediates the association signal. Genomic regions were defined based on LD blocks, and fine-mapping was performed separately for each region. Genes with PIP > 0.9 were considered high-confidence candidates and included in downstream interpretation. FOCUS also incorporates a null model to account for association signals not explained by the tested gene expression models. Similarly, we followed up on brain tissues identified in our tissue-specificity analyses by performing SMR (v1.3.1) analyses, which integrate GWAS and eQTL data in enriched tissues (GTEx v8) to test for causal gene candidates in the prioritised tissues[36]. We applied a Bonferroni multiple-testing correction and accounted for the number of brain tissues in our analysis (0.05 / 5087 [average number of annotations per brain volume] * 13 [number of tested brain tissues] = $7.56 \times 10^{-07}$).

As a conditional post-GWAS analysis, to further understand the influence of BMI in OSA, we sought to identify potential differences in OSA with and without the effect of BMI. Thus, we performed gene-based and gene-set MAGMA (v1.08) and eQTL mapping as implemented in FUMA for $OSA_{mtCOJO\_BMI}$. We implemented a Bonferroni multiple-testing correction, considering the total number of associations (MAGMA gene-based test $P$ = 0.05/18,784 [total number of tested genes] = $2.66 \times 10^{-06}$; eQTL mapping $P$ = 0.05 / 728 [average number of annotations per brain volume] * 13 [number of tested brain tissues] = $5.28 \times 10^{-06}$). We conducted fastBAT analyses with a Bonferroni multiple-testing correction based on the total number of tests ($P$ = 0.05/24,278 (total number of tested genes) = $2.06 \times 10^{-06}$). In addition, we conducted MAGMA's Tissue Expression Analysis using GTEx v8[33] as implemented in FUMA and implemented a Bonferroni multiple-testing correction for general (0.05/30 (total number of general tissues) = $1.67 \times 10^{-03}$) and specific tissues (0.05/54 (total number of specific tissues) = $9.26 \times 10^{-04}$). We also investigated tissue-specific enrichment (for the same tissues tested with the primary OSA meta-analysis) with LDSC-SEG (v1.0.1)[34]. Then, we followed up on brain tissues identified in our tissue-specificity analyses by performing SMR (v1.3.1) analyses. We applied a Bonferroni multiple-testing correction and accounted for the number of brain tissues in our analysis (0.05 / 5,034 [average number of annotations per brain volume] * 13 [number of tested brain tissues] = $7.64x10^{-07}$).

**Genetic correlations**

It is important to note that traits used to estimate genetic correlations were selected based on the following criteria: (i) The relationship between OSA[10,23] and a trait that has been investigated before at a genetic[37,38] or observational[39,40] level, and (ii) The trait has publicly available and well-powered (i.e., large enough sample size to detect genome-wide loci) GWAS summary statistics that can be leveraged to perform LDSC analyses. Thus, GWAS summary data for the other complex human traits included in these analyses were primarily retrieved from international consortia, such as ENIGMA[37,41], the Psychiatric Genetics Consortium (PGC)[42], and the Early Growth Genetics Consortium (EGG)[43], among others.

**Colocalisation**

We leveraged the GWAS-PW (v.0.3.6) method[44] to identify segments of the genome with genetic variants influencing the etiology of OSA and also another human complex trait. For this analysis we leveraged traits with a statistically significant genetic correlation based on our LDSC results to conduct GWAS-PW analyses. Specifically, the GWAS-PW method estimates the posterior probability of association for four different models. In the first model, the segment of the genome is only associated with phenotype A (i.e., OSA in the present study)[44]. For model two, the genomic segment is only influencing the aetiology of phenotype B (i.e., a complex human trait in the present study)[44]. Moreover, in model three, the genomic segment is influencing both phenotypes via the same genetic variants, while in model four the genomic segment influences both phenotypes, but via different genetic variants[44]. In the present study, we provide findings for genomic segments where model three showed a PPA > 0.5, since this threshold has been used in previous studies[37,45–47].

**Potential causal genetic effects**

We selected LHC-MR, as it represents an alternative to traditional Mendelian randomisation methods, which assume no sample overlap between phenotypes[48]. In addition, LHC-MR, unlike traditional Mendelian randomisation methods, leverages the entire set of GWAS summary data (not only genome-wide independent loci) to estimate putative causal genetic effects between two traits. Therefore, LHC-MR has been reported to improve statistical power to estimate bidirectional putative causal genetic effects, direct heritabilities, and confounder effects while accounting for sample overlap[49].

**Polygenic scores estimation and association analyses**

Sleep stage data were originally recorded at 30-second resolution and were down-sampled to 1-minute resolution by assigning each minute the sleep stage recorded during the first 30 seconds. This approach was used to harmonise data for downstream analyses and is unlikely to materially affect results, as sleep stages typically persist for several minutes and Fitbit's proprietary sleep staging algorithm applies temporal smoothing. Sleep features were computed using the filtered 21-day sleep windows and included sleep efficiency (total sleep time divided by total time in bed), WASO (total duration of wake periods following initial sleep onset), number of awakenings (count of discrete wake bouts per night), REM sleep percentage (total REM sleep duration divided by total sleep duration), and fragmentation index (number of epochs classified as wake or light sleep divided by the total number of sleep epochs).
